# Associations of four biological age markers with child development: A multi-omic analysis in the European HELIX cohort

**DOI:** 10.1101/2023.01.23.23284901

**Authors:** Oliver Robinson, ChungHo Lau, Sungyeon Joo, Sandra Andrusaityte, Eva Borràs, Paula de Prado-Bert, Lida Chatzi, Hector C. Keun, Regina Grazuleviciene, Kristine B. Gützkow, Léa Maitre, Dries S. Martens, Eduard Sabido, Valérie Siroux, Jose Urquiza, Marina Vafeiadi, John Wright, Tim Nawrot, Mariona Bustamante, Martine Vrijheid

## Abstract

**Background:** While biological age in adults is often understood as representing general health and resilience, the conceptual interpretation of accelerated biological age in children and its relationship to development remains unclear. We aimed to clarify the relationship of accelerated biological age, assessed through telomere length and three omics-derived biological clocks, to child developmental outcomes, including growth and adiposity, cognition, behaviour, lung function and onset of puberty, among European school-age children participating in the HELIX exposome cohort.

**Methods:** The study population included up to 1,173 children, aged between 5 and 12 years, from study centres in the UK, France, Spain, Norway, Lithuania, and Greece. Telomere length was measured through qPCR, blood DNA methylation and gene expression was measured using microarray, and proteins and metabolites were measured by a range of targeted assays. DNA methylation age was assessed using Horvath’s skin and blood clock, while novel blood transcriptome and “immunometabolic” (based on plasma protein and urinary and serum metabolite data) clocks were derived and tested in a subset of children assessed six months after the main follow-up visit. Associations between biological age indicators with child developmental measures as well as health risk factors were estimated using linear regression, adjusted for chronological age, sex, ethnicity and study centre. The clock derived markers were expressed as Δ age (i.e., predicted minus chronological age).

**Results:** Transcriptome and immunometabolic clocks predicted chronological age well in the test set (r= 0.93 and r= 0.84 respectively). Generally, weak correlations were observed, after adjustment for chronological age, between the biological age indicators. Higher birthweight was associated with greater immunometabolic Δ age, smoke exposure with greater DNA methylation Δ age and high family affluence with longer telomere length. All biological age markers were positively associated with BMI and fat mass, and all markers except telomere length were associated with height, at least at nominal significance (p<0.05). Immunometabolic Δ age was associated with better working memory (p = 4e - 3) and reduced inattentiveness (p= 4e -4), while DNA methylation Δ age was associated with greater inattentiveness (p=0.03) and poorer externalizing behaviours (p= 0.01). Shorter telomere length was also associated with poorer externalizing behaviours (p=0.03).

**Conclusions:** In children, as in adults, biological ageing appears to be a multi-faceted process and adiposity is an important correlate of accelerated biological ageing. Patterns of associations suggested that accelerated immunometabolic age may represent build-up of biological capital while accelerated DNA methylation age and telomere attrition may represent a “wear and tear” model of biological ageing in children.

**Funding:** UK Research and Innovation (MR/S03532X/1); European Commission (grant agreement numbers: 308333; 874583)

## Introduction

The field of geroscience proposes that biological ageing, a set of interrelated molecular and cellular changes associated with ageing, drive the physiological deterioration that is the root of multiple age-related health conditions [1]. Understanding the process of biological ageing and developing markers to accurately assess biological age in individuals, holds great promise for public health and biomedical research in general to develop interventions, even in childhood and early life, that slow physiological decline and reduce the risk of chronic disease and disability in later life.

Telomere length, which shortens with age, is one of the most widely applied biological age markers primarily as it directly assesses a primary Hallmark of Ageing [2, 3]. More recently, high-throughput ‘omic’ methods, which provide simultaneous quantification of thousands of epigenetic marks, transcripts, proteins and metabolites, have been used to develop ‘biological clocks’ that provide a global measure of changes with age at the molecular level [4]. While biological clocks have been primarily trained on chronological age, “age acceleration”, commonly defined as the difference between clock-predicted age and chronological age, has been associated with age-related phenotypes and mortality [5-11], indicating their utility as biological age markers. DNA methylation-based clocks, such as the clock of Horvath [12], have been extensively applied in large-scale studies and remain a research field under active development [13, 14]. Further clocks have been developed using transcriptome [8], metabolome [15] and proteome [9] data, including those that specifically target immune-system related proteins [16]. Generally, clocks have been found to be only weakly correlated with each other, suggesting that each clock captures different facets of biological ageing [17, 18].

While biological age in adults is intuitively understood as an overall indicator of general health and resilience, the conceptual interpretation of biological age acceleration in children is much less clear. Child development and ageing may at first be considered opposing processes, representing growth and decay respectively. However, various related theoretical frameworks link the two processes: Under the developmental origin of health and disease hypothesis, the early life environment is a key determinant of ageing trajectories and disease risk in later life. Life-course models of ageing, supported by measures of physical and cognitive capability, view the childhood developmental phase as key to building up “biological capital” and to determining how long capabilities and disease risk remain above critical thresholds in later years following the gradual decline phase of adult life [19]. Horvath’s DNA methylation clock is currently the only clock trained to predict age throughout the lifespan, and many of the clock’s CpGs are in genomic regions known to regulate development and differentiation [12]. However, unlike life-course models of physical function, the level of DNA methylation at the clock’s CpGs changes in a predictable, unidirectional manner throughout the life-course, albeit at a much faster rate during childhood. This continuous molecular readout suggests that processes directing development are at least indirectly related to detrimental process in later-life and is consistent with quasi-programmed theories of ageing such as antagonistic pleiotropy [20], whereby molecular functions that promote development, inadvertently lead to ageing in later life [21]. Therefore, some authors have suggested that DNA methylation-based age acceleration may be beneficial during childhood [21, 22], reflecting greater physical maturity and build-up of biological capital.

Biological ageing is conceived as continuous balance of cellular damage, caused by both extrinsic environmental factors and by normal physiological processes, and resiliency mechanisms that protect against and compensate for this damage [23]. An alternative “wear and tear” model would view cellular damage to occur continuously from birth and, since the epigenetic clock has been proposed to reflect the epigenomic maintenance system, a resiliency mechanism, DNA methylation age acceleration in children may, as in adults, represent a greater accumulation of epigenetic instability and therefore reduced biological capital. However so far only a handful of studies have examined associations with developmental maturity in children [24-26]. Telomere length attrition is more rapid in early childhood during rapid somatic growth and more gradual in adulthood, with those with a shorter telomere length in childhood maintaining a lower telomere length into adulthood [27]. While telomere length may serve as both a mitotic-clock and as a mediator of cellular stress [28], the associations reported between environmental stress in childhood and shorter telomere length suggest it reflects early-life cellular damage that may be carried into adulthood.

Little is known regarding the interpretation of biological age in children assessed at the transcriptome, proteomic and metabolomic levels, since few biological clocks are available for this age range using these data. To the best of our knowledge, only the study of Giallourou et al [29] has applied metabolomic data to provide a multivariate model of age in children, finding that growth constrained infants lag in their metabolic maturity relative to their healthier peers. It is possible that biological clocks constructed using these data, particularly proteomic and metabolomics, support the life-course ageing framework, where age acceleration in children represents a buildup of biological capital, since they are closer to the phenotype than the DNA-based epigenetic clocks and telomere length.

To explore these questions, we have performed a comparative analysis of four assessments of biological age within the pan-European Human Early Life Exposome (HELIX) cohort of children aged between 5 and 12 years. We hypothesized that biological age measures would be associated with child development. We systematically compared associations with developmental endpoints, including growth and adiposity, cognition, behaviour, lung function and pubertal development, and common health risk factors, for telomere length, DNA methylation age, and two newly derived clocks: transcriptome age and immunometabolic age. Through this analysis, we aimed to clarify the relationship of age acceleration in children to the buildup of biological capital, and more broadly develop new biological markers of overall developmental staging in children.

## Results

### Sample characteristics

We used blood or urine derived measurements from the pan-European HELIX cohort. This included blood telomere length (*N =* 1,162), blood DNA methylation (*N* = 1,173, 450K CpGs), blood gene expression (N = 1,007, 50K genes), and proteins and metabolites (N= 1,152, 36 plasma proteins, 177 serum metabolites and 44 urinary metabolites), with 869 children overlapping across all measurements. Each subsample included around 55% boys, 89% children of white European ancestry, and a mean age of around 8 years (range 5-12 years). Around 51% of mothers of the HELIX children in each subsample had a high education level. The HELIX cohort included children from six study centres based in the UK, Spain, Greece, Lithuania, France, and Norway, with each centre contributing between 11 and 24% to each subsample (see Table 1 for sample characteristics).

**Table 1.** Summary Statistics for study population.

|  | TELOMERE<br>LENGTH | DNA<br>METHYLATION<br>AGE | TRANSCRIPT-<br>OME AGE | IMMUNO-METABOLIC AGE |
| --- | --- | --- | --- | --- |
| N | N (%) or<br>Mean (SD)<br>1,162 | N (%) or Mean<br>(SD)<br>1,173 | N (%) or<br>Mean (SD)<br>1,007 | N (%) or Mean<br>(SD)<br>1,152 |
| **DEMOGRAPHIC FACTORS** |  |  |  |  |
| **AGE (YEARS)** | 7.84 (1.54) | 7.84 (1.54) | 7.90(1.50) | 7.86 (1.55) |
| **SEX-MALE** | 639 (55) | 644 (54.9) | 547 (54.3) | 628 (54.5) |
| **SEX-FEMALE** | 523 (45) | 529 (45.1) | 460 (45.7) | 524 (45.5) |
| **ETHNICITY-WHITE** | 1039 (89.4) | 1048 (89.3) | 905 (89.9) | 1032 (89.6) |
| **ETHNICITY-PAKISTANI/ASIAN** | 96 (8.3) | 98 (8.4) | 76 (7.5) | 93 (8.1) |
| **ETHNICITY -OTHER** | 27 (2.3) | 27 (2.3) | 26 (2.6) | 27 (2.3) |
| **COHORT-BIB** | 200 (17.2) | 203 (17.3) | 162 (16.1) | 191 (16.6) |
| **COHORT-EDEN** | 145 (12.5) | 146 (12.4) | 109 (10.8) | 149 (12.9) |
| **COHORT-INMA** | 212 (18.2) | 215 (18.3) | 184 (18.3) | 201 (17.4) |
| **COHORT-KANC** | 196 (16.9) | 198 (16.9) | 151 (15) | 197 (17.1) |
| **COHORT-MOBA** | 211 (18.2) | 212 (18.1) | 245 (24.3) | 222 (19.3) |
| **COHORT-RHEA** | 198 (17) | 199 (17) | 156 (15.5) | 192 (16.7) |

| PRENATAL FACTORS |  |  |  |  |
| --- | --- | --- | --- | --- |
| MATERNAL NON-ACTIVE SMOKER DURING PREGNANCY | 988 (85) | 998 (85.1) | 859 (85.3) | 981 (85.2) |
| MATERNAL ACTIVE SMOKER DURING PREGNANCY | 174 (15) | 175 (14.9) | 148 (14.7) | 171 (14.8) |
| BIRTHWEIGHT (KG) | 3.37 (0.5) | 3.37 (0.5) | 3.38 (0.52) | 3.38 (0.5) |
| GESTATIONAL AGE (WEEKS) | 39.57 (1.67) | 39.58 (1.67) | 39.59 (1.75) | 39.59 (1.66) |
| FAMILY CAPITAL |  |  |  |  |
| MATERNAL EDUCATION (LOW) | 165 (14.7) | 166 (14.7) | 140 (14.4) | 157 (14.1) |
| MATERNAL EDUCATION (MEDIUM) | 391 (34.8) | 394 (34.8) | 328 (33.8) | 391 (35.1) |
| MATERNAL EDUCATION (HIGH) | 568 (50.5) | 573 (50.6) | 503 (51.8) | 565 (50.8) |
| FAMILY AFFLUENCE (LOW) | 133 (11.5) | 135 (11.5) | 112 (11.1) | 128 (11.1) |
| FAMILY AFFLUENCE (MEDIUM) | 462 (39.8) | 466 (39.8) | 394 (39.2) | 450 (39.1) |
| FAMILY AFFLUENCE (HIGH) | 565 (48.7) | 570 (48.7) | 499 (49.7) | 572 (49.7) |
| FAMILY SOCIAL CAPITAL (LOW) | 513 (47.7) | 516 (47.5) | 422 (45.8) | 496 (46.7) |
| FAMILY SOCIAL CAPITAL (MEDIUM) | 264 (24.6) | 269 (24.8) | 228 (24.7) | 259 (24.4) |
| FAMILY SOCIAL CAPITAL (HIGH) | 298 (27.7) | 301 (27.7) | 272 (29.5) | 307 (28.9) |
| CHILD FACTORS |  |  |  |  |
| NO PASSIVE SMOKE EXPOSURE | 723 (63.8) | 732 (63.9) | 639 (64.5) | 718 (63.8) |
| PASSIVE SMOKE EXPOSURE | 411 (36.2) | 413 (36.1) | 351 (35.5) | 407 (36.2) |
| PHYSICAL ACTIVITY-LOW | 418 (36.9) | 420 (36.8) | 349 (35.3) | 416 (37.1) |
| PHYSICAL ACTIVITY-MEDIUM | 336 (29.7) | 341 (29.9) | 295 (29.9) | 330 (29.4) |
| PHYSICAL ACTIVITY-HIGH | 378 (33.4) | 381 (33.4) | 344 (34.8) | 375 (33.5) |
| KIDMED DIET SCORE | 2.81 (1.77) | 2.82 (1.78) | 2.88 (1.77) | 2.84 (1.76) |
| DEVELOPMENTAL MEASURES |  |  |  |  |
| HEIGHT Z-SCORE | 0.4 (0.97) | 0.39 (0.98) | 0.39 (0.96) | 0.4 (0.98) |
| BMI Z-SCORE | 0.43 (1.2) | 0.43 (1.2) | 0.4 (1.15) | 0.42 (1.18) |
| ADIPOSIITY (BIA FAT-MASS %) | 6.76 (4.01) | 6.77 (4.01) | 6.52 (3.9) | 6.72 (3.95) |
| WORKING MEMORY (3-BACK D') | 1.1 (1.01) | 1.1 (1.01) | 1.13 (1) | 1.1 (1.01) |
| INATTENTIVENESS (ANT-HRT) | 301.97 (90.38) | 301.93 (90.46) | 297.69 (89.36) | 301.35 (89.84) |
| FLUID INTELLIGENCE (CPM) | 25.87 (6.33) | 25.86 (6.32) | 26.12 (6.26) | 25.95 (6.3) |
| INTERNALIZING BEHAVIOURS (CBCL) | 6.49 (5.9) | 6.48 (5.9) | 6.36 (5.89) | 6.52 (5.87) |
| EXTERNALIZING BEHAVIOURS (CBCL) | 6.81 (6.5) | 6.82 (6.51) | 6.67 (6.49) | 6.74 (6.42) |
| LUNG FUNCTION (FEV1) | 99.26 (13.46) | 99.25 (13.47) | 99.16 (13.02) | 99.17 (13.47) |
| PUBERTY NOT STARTED | 250 (46.6) | 252 (46.5) | 254 (49.7) | 260 (48) |
| PUBERTY STARTED (PDS >1) | 287 (53.4) | 290 (53.5) | 257 (50.3) | 282 (52) |

### Biological age marker performance

We assessed four markers of biological age: telomere length, DNA methylation age, transcriptome age and ‘immunometabolic’ age (Figure 1). DNA methylation age was calculated using the published Skin and blood Horvath clock [30] to allow greater comparison to the wider literature, including in adults. We previously reported this epigenetic clock to show the best performance in chronological age prediction within the HELIX cohort [31]. Since no published applicable transcriptome, proteome or metabolome clocks were available for the age range of our sample, we trained two new biological clocks using these data in the HELIX cohort, through elastic net regression and cross-validation. We combined the proteome and metabolome data into a single immunometabolic age clock, since the available proteomic data included biomarkers targeting both metabolic and inflammatory functions, both omic types represent final products of gene regulation, and since the metabolic and immune systems are closely linked [32].

**Figure 1:**
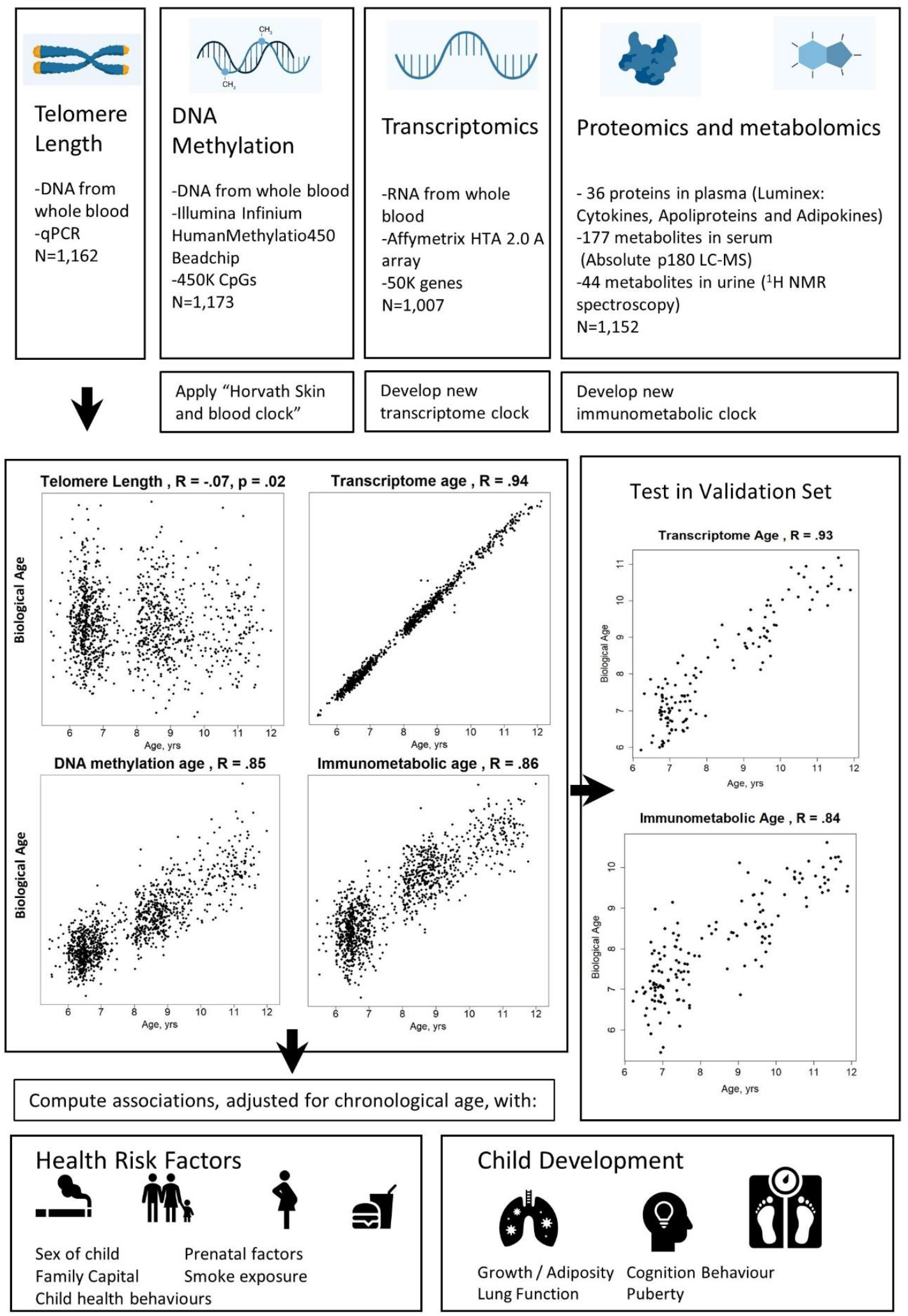
Study design schematic.

The correlation between telomere length and chronological age was weak but statistically significant (r = -0.07, p= 0.02). Correlations with chronological age were r= 0.85 for DNA methylation age, r= 0.94 for transcriptome age, and r = 0.86 for immunometabolic age (Figure 1).

We validated the transcriptome and immunometabolic clocks using cross-validation within the HELIX subcohort (cross-validated r of 0.87 and 0.82 respectively) and further tested in a subset of children who attended a second clinic visit approximately 0.5 years after the main follow-up visit (standard deviation (SD) = 0.18 years) as part of the HELIX panel study. Correlations in this test set were r= 0.93 for transcriptome age (N= 128) and r= 0.84 for immunometabolic age (N=151) (Figure 1). Predicted biological age increased by mean 0.33 years (SD =0.58) for transcriptome age (t-test, p=3e -5) and mean 0.22 years (SD: 0.59 years) for immunometabolic age (t-test, p=2e -5) between the first and second visits (Figure S1). Correlations were significant (p < 0.05) within each study centre for both clocks, except for immunometabolic age for children from the BiB (UK) cohort (Figures S2 and S3).

The immunometabolic age clock was composed of 135 predictors including 20 proteins, 79 serum metabolites and 36 urinary metabolites (table S1). The transcriptome clock was composed of 1,445 genes, 652 of which were annotated to Gene Symbols (table S2). The transcriptome clock genes were enriched (false discovery rate (FDR)-corrected p < 0.05) in ‘ribosome’ and ‘ribosome biogenesis’ KEGG pathways (table S3) and the following level 2 Gene Ontology biological process terms: ‘leukocyte activation’, ‘movement of cell or subcellular component’, ‘leukocyte migration’, ‘cell activation’, and ‘secretion by cell’ (table S4). We also tested enrichment of transcriptome clock predictors among genes reported by a large meta-analysis of age in adults [8]: among the 1,406 reported age-associated genes that could be matched to our measured genes, 43 were included in our transcriptome clock (hypergeometric enrichment test, p = 0.052).

Figure 2 shows partial correlations, adjusted for chronological age and study centre, between the biological age markers. Only null to weak correlations were observed, with significant correlations between telomere length and DNA methylation age (r = -0.06, p = 0.04) and between transcriptome age and immunometabolic age (r = 0.08, p = 0.01).

**Figure 2:**
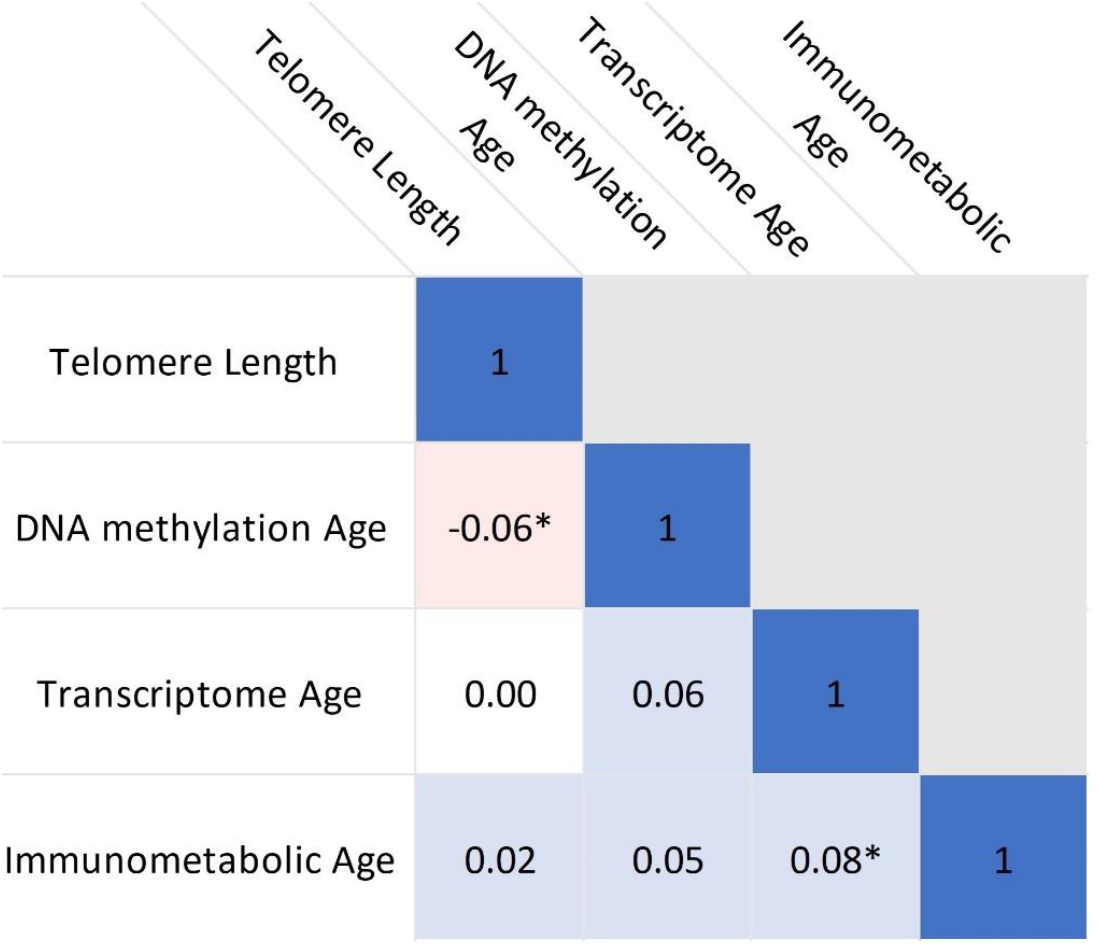
Correlations between biological age indicators. Heatmap shows partial Pearson’s correlations, adjusted for chronological age, sex and study centre. * indicates p<0.05.

### Biological clock associations with health risk factors

Table 2 shows associations, adjusted for chronological age, sex, study centre and ethnicity, between health risk factors and the biological age markers. The markers derived from omic-based biological clocks are expressed as Δ age (clock-predicted age – chronological age) and since the adjustment set included chronological age, effects can be interpreted as years of age acceleration as often defined [18].

**Table2:**
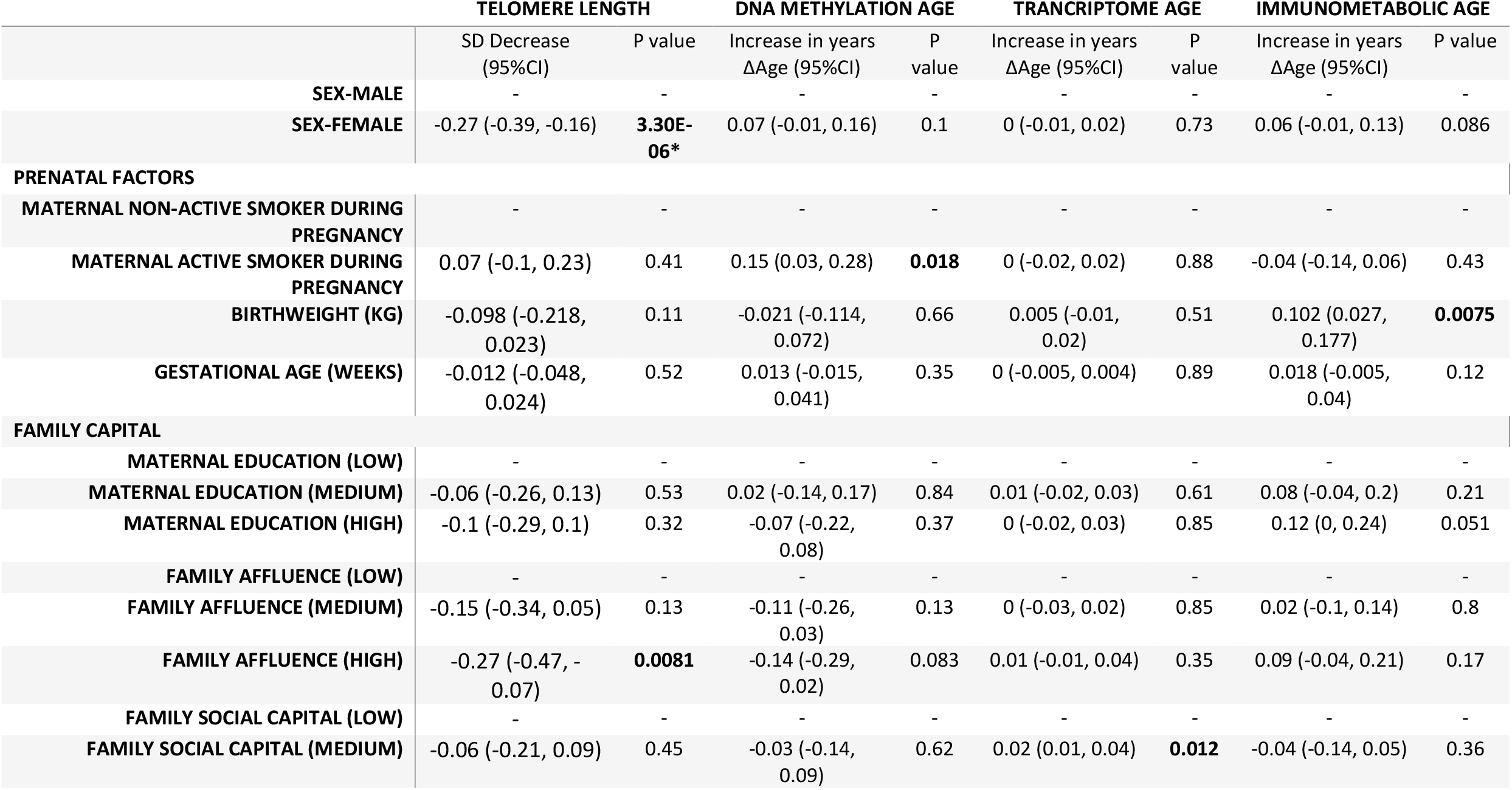
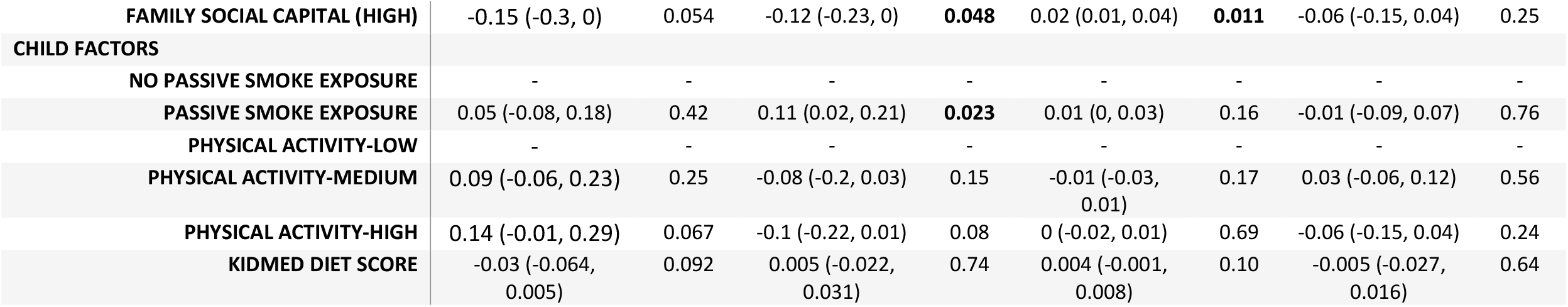
Associations between health risk factors and biological age measures. Estimates calculated using linear regression, adjusted for chronological age, sex, ethnicity, and study centre. Bold indicates p<0.05 and *indicates FDR <5%. Telomere length is expressed as % decrease in length (multiplied by -1) to provide estimates indicative of accelerated biological age, as the other biological age indicators.

|  | TELOMERE LENGTH |  | DNA METHYLATION AGE |  | TRANSCRIPTOME AGE |  | IMMUNOMETABOLIC AGE |  |
| --- | --- | --- | --- | --- | --- | --- | --- | --- |
| | SD Decrease<br>(95%CI) | P value | Increase in years<br>ΔAge (95%CI) | P<br>value | Increase in years<br>ΔAge (95%CI) | P<br>value | Increase in years<br>ΔAge (95%CI) | P value |
| SEX-MALE | - | - | - | - | - | - | - | - |
| SEX-FEMALE | -0.27 (-0.39, -0.16) | **3.30E-06\*** | 0.07 (-0.01, 0.16) | 0.1 | 0 (-0.01, 0.02) | 0.73 | 0.06 (-0.01, 0.13) | 0.086 |
| **PRENATAL FACTORS** |  |  |  |  |  |  |  |  |
| MATERNAL NON-ACTIVE SMOKER DURING PREGNANCY | - | - | - | - | - | - | - | - |
| MATERNAL ACTIVE SMOKER DURING PREGNANCY | 0.07 (-0.1, 0.23) | 0.41 | 0.15 (0.03, 0.28) | **0.018** | 0 (-0.02, 0.02) | 0.88 | -0.04 (-0.14, 0.06) | 0.43 |
| BIRTHWEIGHT (KG) | -0.098 (-0.218, 0.023) | 0.11 | -0.021 (-0.114, 0.072) | 0.66 | 0.005 (-0.01, 0.02) | 0.51 | 0.102 (0.027, 0.177) | **0.0075** |
| GESTATIONAL AGE (WEEKS) | -0.012 (-0.048, 0.024) | 0.52 | 0.013 (-0.015, 0.041) | 0.35 | 0 (-0.005, 0.004) | 0.89 | 0.018 (-0.005, 0.04) | 0.12 |
| **FAMILY CAPITAL** |  |  |  |  |  |  |  |  |
| MATERNAL EDUCATION (LOW) | - | - | - | - | - | - | - | - |
| MATERNAL EDUCATION (MEDIUM) | -0.06 (-0.26, 0.13) | 0.53 | 0.02 (-0.14, 0.17) | 0.84 | 0.01 (-0.02, 0.03) | 0.61 | 0.08 (-0.04, 0.2) | 0.21 |
| MATERNAL EDUCATION (HIGH) | -0.1 (-0.29, 0.1) | 0.32 | -0.07 (-0.22, 0.08) | 0.37 | 0 (-0.02, 0.03) | 0.85 | 0.12 (0, 0.24) | 0.051 |
| FAMILY AFFLUENCE (LOW) | - | - | - | - | - | - | - | - |
| FAMILY AFFLUENCE (MEDIUM) | -0.15 (-0.34, 0.05) | 0.13 | -0.11 (-0.26, 0.03) | 0.13 | 0 (-0.03, 0.02) | 0.85 | 0.02 (-0.1, 0.14) | 0.8 |
| FAMILY AFFLUENCE (HIGH) | -0.27 (-0.47, -0.07) | **0.0081** | -0.14 (-0.29, 0.02) | 0.083 | 0.01 (-0.01, 0.04) | 0.35 | 0.09 (-0.04, 0.21) | 0.17 |
| FAMILY SOCIAL CAPITAL (LOW) | - | - | - | - | - | - | - | - |
| FAMILY SOCIAL CAPITAL (MEDIUM) | -0.06 (-0.21, 0.09) | 0.45 | -0.03 (-0.14, 0.09) | 0.62 | 0.02 (0.01, 0.04) | **0.012** | -0.04 (-0.14, 0.05) | 0.36 |
| **FAMILY SOCIAL CAPITAL (HIGH)** | -0.15 (-0.3, 0) | 0.054 | -0.12 (-0.23, 0) | **0.048** | 0.02 (0.01, 0.04) | **0.011** | -0.06 (-0.15, 0.04) | 0.25 |
| **CHILD FACTORS** |  |  |  |  |  |  |  |  |
| **NO PASSIVE SMOKE EXPOSURE** | - | - | - | - | - | - | - | - |
| **PASSIVE SMOKE EXPOSURE** | 0.05 (-0.08, 0.18) | 0.42 | 0.11 (0.02, 0.21) | **0.023** | 0.01 (0, 0.03) | 0.16 | -0.01 (-0.09, 0.07) | 0.76 |
| **PHYSICAL ACTIVITY-LOW** | - | - | - | - | - | - | - | - |
| **PHYSICAL ACTIVITY-MEDIUM** | 0.09 (-0.06, 0.23) | 0.25 | -0.08 (-0.2, 0.03) | 0.15 | -0.01 (-0.03, 0.01) | 0.17 | 0.03 (-0.06, 0.12) | 0.56 |
| **PHYSICAL ACTIVITY-HIGH** | 0.14 (-0.01, 0.29) | 0.067 | -0.1 (-0.22, 0.01) | 0.08 | 0 (-0.02, 0.01) | 0.69 | -0.06 (-0.15, 0.04) | 0.24 |
| **KIDMED DIET SCORE** | -0.03 (-0.064, 0.005) | 0.092 | 0.005 (-0.022, 0.031) | 0.74 | 0.004 (-0.001, 0.008) | 0.10 | -0.005 (-0.027, 0.016) | 0.64 |

Nominally significant associations were observed for the following health risk factors: Telomere length was longer among girls compared to boys (p**=** 3e -06) and among children of high affluence families (p= 0.008). DNA methylation Δ age was higher among children of mothers who actively smoked during pregnancy (p= 0.018) and children exposed to passive smoke (p= 0.023), while DNA methylation Δ age was lower among children from families with high social capital (p= 0.048). Conversely, transcriptome Δ age was positively associated with medium and high (p= 0.011) family social capital. Immunometabolic Δ age was associated with higher birthweight (p= 0.0075). Only the association between longer telomere length and female sex passed FDR correction

### Biological age associations with development

Figure 3 and table S5 shows associations, adjusted for chronological age, sex, study centre and ethnicity, between the biological age markers and developmental outcomes related to growth and adiposity, cognition, behaviour, lung function and onset of puberty.

**Figure 3:**
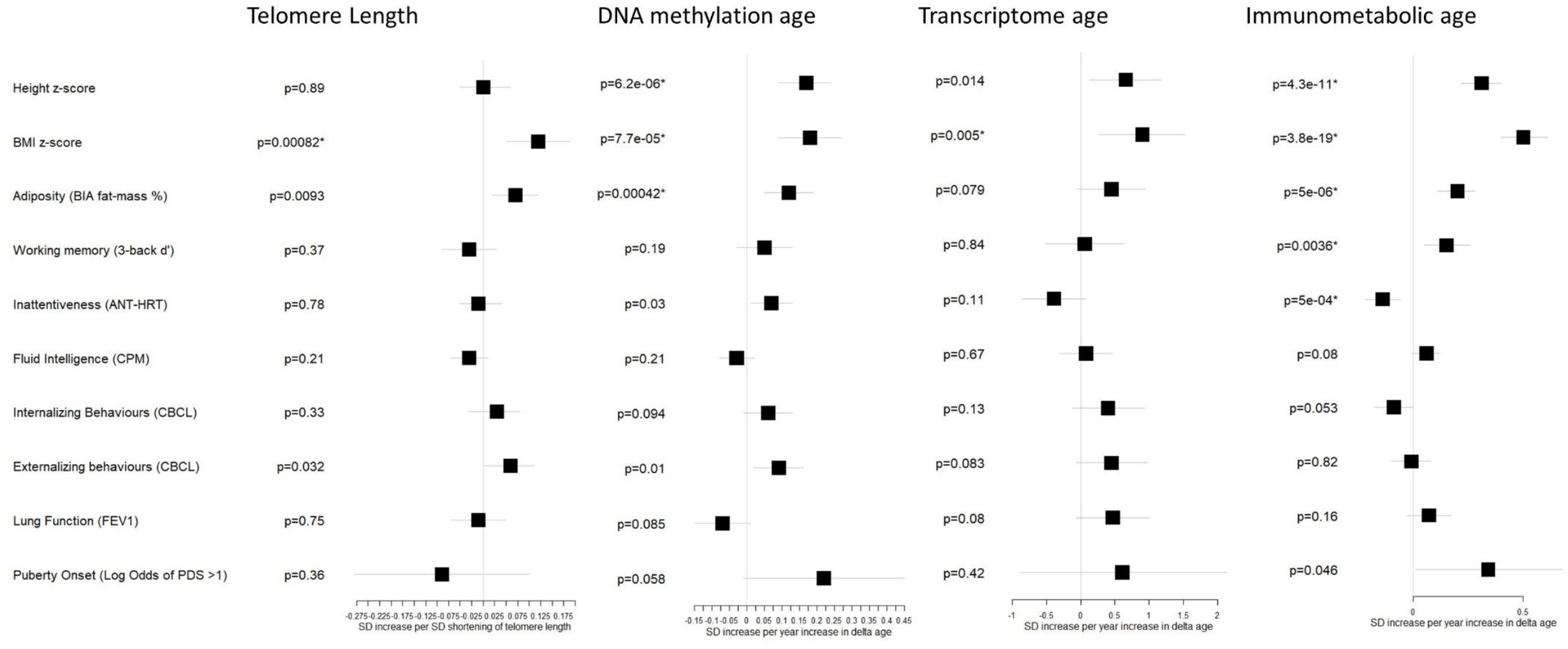
Associations between biological age measures and developmental measures. Estimates calculated using linear regression, adjusted for chronological age, sex, ethnicity, and study centre. *indicates FDR <5%. Telomere length is expressed as % decrease in length (multiplied by -1) to provide estimates indicative of accelerated biological age, as the other biological age indicators. SD = Standard deviation

Several developmental outcomes were associated with biological age markers after FDR correction: DNA methylation and immunometabolic Δ age were associated with greater height z-score (p= 6e -6 and p= 4e -11 respectively) and greater fat mass % (p= 0.0004 and p= 5e -6 respectively). All biological age markers were associated with greater BMI z-score (telomere length p =8e -4, DNA methylation Δ age p = 8e -5, transcriptome Δ age p = 0.005, immunometabolic Δ age p =4e -19). Furthermore, immunometabolic Δ age was associated after FDR correction with better working memory (p = 0.0036) and reduced inattentiveness (p= 5e -4).

Associations at the nominal significance (p<0.05) level were observed for increases in height z-score with transcriptome Δ age (p= 0.014), shorter telomere length with increased fat mass % (p= 0.009), and DNA methylation Δ age with greater inattentiveness (p=0.03). Both shorter telomere length and DNA methylation Δ age were associated with greater externalizing behaviours (p= 0.032 and p= 0.01 respectively). Among a smaller subset of children (table 1) aged over 8 years, we observed a nominally significant association between immunometabolic Δ age and odds of onset of puberty (Odds Ratio: 1.41, 95% CI: 1.01, 1.97, p =0.046).

No significant associations with lung function were observed, but like the patterns of associations observed with cognitive and behavioural outcomes, there was a trend for a negative association with DNA methylation Δ age (p = 0.085) and a positive association with immunometabolic Δ age (p=0.16).

### Sensitivity Analysis

In sensitivity analysis. We firstly stratified by sex and generally observed similar associations among boys and girls, apart from the following differences (Figure S4): Associations between shorter telomere length and BMI z-score and adiposity were stronger among boys. For DNA methylation Δ age, associations with poorer externalizing and internalizing behaviours were only apparent among boys. For transcriptome Δ age, stronger associations among boys were observed with BMI z-score, adiposity and poorer externalizing and internalizing behaviours. Conversely, we observed an association between transcriptome Δ age and reduced inattentiveness among girls only. Immunometabolic Δ age was more strongly associated with reduced inattentiveness among girls and also associated with greater odds of puberty onset among girls only.

Secondly, we additionally adjusted our models by estimated cell counts to determine the influence of cell composition on associations with developmental outcomes (Figures S5B, S6B, S7B and S8B, Table S5). Associations were generally little changed: For DNA methylation Δ age, associations were attenuated with adiposity and growth outcomes although all remained FDR significant and the association with externalizing behavior was slightly attenuated. For transcriptome Δ age associations with adiposity and growth outcomes and lung function increased slightly and the association with greater lung function became nominally significant.

Finally, we assessed the effects of further adjustment for health risk factors (family affluence and social capital, birthweight, maternal active smoking, and child passive smoking) since health risk factors could be independently associated with both biological age and developmental outcomes (Figures S5C, S6C, S7C and S8C, Table S5). Associations were generally little changed, expect for an attenuation of the association between telomere length and externalizing behavior, while conversely the association between DNA methylation Δ age and externalizing behavior was slightly strengthened.

## Discussion

In a large sample of European children, we have analysed four measures of biological age, derived from molecular features at different levels of biological organization, in relation to developmental outcomes and health risk factors. We assessed two established biological age markers, telomere length and DNA methylation age, and derived two new measures, transcriptome age and immunometabolic age. Despite finding only null to weak correlations between the measures, we found all measures to be positively associated with greater BMI and adiposity, and both DNA methylation Δ age and immunometabolic Δ age were associated with taller height. While immunometabolic Δ age was associated with greater cognitive maturity including greater working memory and attentiveness, conversely DNA methylation Δ age was nominally associated with greater inattentiveness and both DNA methylation Δ age and shorter telomere length were nominally associated with poorer externalizing behaviours.

BMI has consistently been associated with accelerated ageing in adults across a diverse range of biological age markers [8, 18, 33, 34] underlining the integral role of metabolism in ageing. Indeed, a recent large study of Dutch adults found BMI to be the only health risk factor tested associated with accelerated ageing across five biological age clocks, including telomere length, DNA methylation, transcriptome, proteome and metabolomic age markers [18]. Here we show that the link between BMI and multiple dimensions of accelerated ageing is also apparent in children. Energy and nutrient intake influence all Ageing Hallmarks and multiple lines of evidence link increased adiposity to shorter lifespan [35]. These effects appear to be partially mediated through evolutionarily conserved nutrient sensing systems such as the mTOR signaling pathway, which promote anti-ageing cellular repair mechanisms, at the expense of growth and metabolism, in response to lower nutrient availability [35]. Furthermore, excess adiposity increases generalized inflammation and oxidative stress [36, 37], which may have direct effects on age markers, particularly telomere length and DNA methylation age acceleration.

The observed associations between greater height with biological age may indicate developmental maturity. Height is generally considered reflective of a beneficial early-life environment [38], however, evidence for an association with lifespan is mixed [38, 39], with a recent meta-analysis suggesting a u-shaped relationship with all-cause mortality [40]. Greater comparative height at 10 years was also inversely associated with longevity in a recent large-scale Medelian randomization study [41]. Furthermore, there is some evidence that the link between height and longevity may be mediated through the insulin-like growth factor-1 signaling pathway [39, 42]. The associations may also be interpreted as greater rates of growth and anabolism exerting greater “wear and tear” on cellular structures. Two other studies have also observed an association between height and DNA methylation age acceleration in children [25, 26].

Despite similarities in associations with growth and adiposity measures, patterns of association across cognitive and behavioral domains varied across biological age markers, underlying the view of biological ageing as a multi-faceted process. Immunometabolic Δ age was associated with greater cognitive maturity, fitting the life-course model of greater accumulation of biological capital during the build-up phase of development. Immunometabolic Δ age may be considered a phenotypic summary measure of metabolic and immune system maturity, and these cognitive developmental associations suggest that it may also be generalisable to overall developmental stage. On the other hand, DNA methylation Δ age was related to relative immaturity in attentiveness and externalizing behaviour. A previous Finish study of children age between 11 and 13 years also reported associations between DNA methylation age acceleration and behavioural problems [26]. Similarly, shorter telomere length was associated here with greater externalizing behaviours, although not with any cognitive domains. Four other studies have examined the link between shorter telomere length and externalizing behaviours [43-46], with all, except one [46], also reporting an association.

Overall patterns of associations between risk factors and biological age measures also suggest the detrimental nature of accelerated ageing in children assessed through telomere length and DNA methylation Δ age, and potentially beneficial nature of advanced immunometabolic Δ age. Both prenatal maternal active smoking and child passive smoking were associated with DNA methylation Δ age, while greater birthweight was associated with immunometabolic Δ age. We examined maternal education level, family affluence, and social capital that broadly represent the three forms of interconvertible capital (cultural, economic, and social) proposed by Bourdieu [47]. It has been theorized that biological capital represents a fourth type of human capital, and that the conversion across these forms of capital underlies inequalities in ageing trajectories [48]. Nominally significant associations between higher family affluence with longer telomere length and high social capital with a younger DNA methylation age indicate that age acceleration assessed through these measures does not represent an accumulation of biological capital. Generally, directions of effect for immunometabolic Δ age were in the opposite direction which may suggest it represents greater biological capital.

Girls were found to have longer telomere lengths than boys. Women have been consistently found to have longer telomere lengths [49] although the few generally smaller studies in children have been inconsistent [45, 50-52]. No other biological age markers were associated with sex, which contrasts with the study of Jansen et al [18] in adults which reported accelerated biological age in men across all measures tested except for proteomic age. Indeed, the phenomenon of accelerated DNA methylation age in men is well-established [21], consistent with lower life-expectancies for men. Although it is not known if these biological age differences are due to biological mechanisms or greater prevalence of disease risk factors among men, our data in children before divergence of risk factor prevalence could indicate a biological mechanism for telomere sex differences and a risk factor mediated mechanism for other biological age markers. Interestingly, we observed differences in associations between biological age and development between boys and girls, with some consistency across markers: Both shorter telomere length and transcriptome Δ age and were more strongly associated with adiposity in boys, DNA methylation and transcriptome Δ age showed stronger associations among boys with poorer behaviour, while in girls both transcriptome and immunometabolic Δ age showed stronger associations with improved attentiveness. Given observed sexual dimorphism in both developmental rates [53] and biological age measures through a variety of proposed mechanisms [54], it may be unsurprising that relationship between biological age and development also differs between the sexes.

Furthermore, we observed that immunometabolic Δ age was associated with greater odds of puberty onset, driven by effects observed among girls only. We did not observe any further significant associations with onset of puberty, however the sample size in the subset of children was small compared to the other developmental measures. There was also suggestive evidence for associations between DNA methylation Δ age with onset of puberty with associations close to the nominal significance threshold. Three previous studies have reported associations between DNA methylation age acceleration and puberty onset and stage [24-26], and one study has reported associations between shorter telomere length and puberty onset [55]. However, directions of effect for telomere length in our study were in the opposite direction. While earlier age at puberty is representative of more advanced physical maturation, it has been associated with metabolic diseases in later life, including cancers [56] and all-cause mortality [57].

We found transcriptome data to be highly accurate in predicting chronological age, including in a test set of children assessed six months later, demonstrating that gene expression tracks closely with age in children, even over this relatively short period. We analyzed biological pathways and processes enriched among transcript clusters contributing to the transcriptome clock, observing the integral role of ribosome and ribosome biogenesis pathways, central to protein synthesis, and biological processes including the immunity related processes leukocyte migration and activation, and cell movement, activation, and secretion. Strikingly, gene expression in adults is similarly characterized by downregulation of ribosomal genes and enrichment of expression in immune related genes [8]. This indicates that, similar to DNA methylation changes [21], there is some overlap in gene expression related to both development in children and ageing in adults. Although formal testing of enrichment of genes contributing to transcriptome clock presented here among age-associated genes in adults showed enrichment at only borderline statistical significance, the transcriptome clock predictors are an underrepresentation of the full profile of gene expression associated with age in children, due to the sparsity enforced during the variable selection training process.

Despite associations with growth and adiposity measures, transcriptome age generally showed weaker associations with other developmental outcomes than for the other biological age markers. While this in part can be attributed to the slightly smaller sample size for children with transcriptome data, it is also likely due to the high accuracy in predicting chronological age of the transcriptome clock, resulting in lower variation in the portion of transcriptome age that is not explained by chronological age, further reducing statistical power. This makes it challenging to judge the relevance of transcriptome age, if any, to developmental endpoints, which may be mixed since, non-significant direction of effects were observed with both maturity in attention and lung function, yet relative immaturity in behaviour. In fact, training clocks using chronological age, which while providing an accessible route to understanding molecular changes associated with age, does pose limitations generally for inference regarding biological ageing. Particularly for high-dimensional data such as DNA methylation, it has been shown that it is possible to predict chronological age near-perfectly [58], thereby limiting information on biological age and its variation. For this reason, newer epigenetic clocks have included clinical and mortality data, to improve clinical relevance and sensitivity to risk factors [13, 14], which should be considered in future studies developing clocks in children.

Other limitations include the cross-sectional design of the main analysis, which limits inference regarding the directionality of associations and allows the possibility of age-associated environmental factors to influence the clock development. Furthermore, there were differences in age by study centre. Children from the EDEN cohort study centre were generally older, which likely introduced a degree of cohort bias into the age modelling. For this reason, we adjusted all associations by study centre and additionally assessed age correlation within each study centre. Although cohorts were recruited from the general population, certain ethnicities or socio-economically disadvantaged groups may have been under-represented, limiting generalizability somewhat. A bias towards over-representation of White ethnic groups is an issue generally with the development of biological clocks, which means associations observed with ethnicity should be interpreted cautiously. While the DNA methylation and transcriptome data was representative of the full genome, our coverage of the metabolome and proteome was limited to targeted assays. However, the strengths of this study include the large population sample, drawing from six countries from around Europe, increasing generalizability, and the integration of rich molecular data and a broad range of developmental outcomes into a single systematic analysis. Although our age range was somewhat limited, missing the infancy and adolescent periods, the age range covered a key childhood period, where energy expenditure (an indicator of level of overall physiology) has entered a period of steady increase following more rapid increases during infancy and before stabilization during adolescence [59].

In conclusion, in this large Pan-European study we have found that four indicators of biological age, representing complimentary molecular processes, were all associated with BMI after controlling for chronological age, indicating that adiposity is an important correlate of accelerated biological ageing in children. We developed a highly accurate “transcriptome age” clock although it was found to be relatively insensitive to other development phenotypes. We found that immunometabolic Δ age was associated with cognitive maturity fitting a buildup of biological capital model of ageing in children, while shorter telomere length and DNA methylation Δ age was associated with greater behavioral problems suggesting a “wear and tear” model of ageing in children. Our findings contribute to the interpretation and understanding of biological age measures in children, crucial for clinical and epidemiological research into early life risk factors for adverse ageing trajectories. Future long-term studies should investigate associations between age acceleration in children and adults to further test the antagonistic pleiotropy hypothesis.

## Materials and Methods

### Study population

This study population included children recruited into the European population-based HELIX exposome cohort [60, 61], which was based on six on-going longitudinal population-based birth cohorts established in six countries across different parts of Europe (Born in Bradford [BiB; UK] [62], Étude des Déterminants Pré et Postnatals du Développement et de la Santé de l’Enfant [EDEN; France] [63], Infancia y Medio Ambiente [INMA; Spain] [64], Kaunas Cohort [KANC; Lithuania] [65], Norwegian Mother, Father and Child Cohort Study [MoBa; Norway] [66], and Mother-Child Cohort in Crete [RHEA; Greece] [67]) covering singleton deliveries from 2003 to 2008. All children participated in a harmonized ‘HELIX subcohort’ clinical examination in their respective study centres during 2014-2015, where biological samples were collected. A subset of children (from all study centres apart from MoBa), attended a second clinical examination, as part of the ‘HELIX panel study’ approximately 6 months after the first ‘HELIX subcohort’ examination, where a similar suite of biological samples were collected. A full description of the HELIX follow-up methods and study population, including eligibility criteria and sample size calculations are available in [60, 61]. In the current study we included all children with available molecular data (figure S9).

Prior to the start of HELIX, all six cohorts had undergone the required evaluation by national ethics committees and obtained all the required permissions for their cohort recruitment and follow-up visits. Each cohort also confirmed that relevant informed consent and approval were in place for secondary use of data from pre-existing data. The work in HELIX was covered by new ethical approvals in each country and at enrolment in the new follow-up, participants were asked to sign a new informed consent form. Additionally, the current study was approved by the Imperial College Research Ethics Committee (Reference: 19IC5567).

### Biological sample collection and processing

Blood was collected at the end of the clinical examination of the child to ensure an approximate 3 hours (median = 3.5 hours, SD = 1.1 hour) fasting time since the last meal. Blood samples were collected using a ‘butterfly’ vacuum clip and local anaesthetic and processed into a variety of sample matrices, including plasma, whole blood for RNA extraction (Tempus tubes - Life Technologies, USA), red cells, and buffy coat for DNA extraction. These samples were frozen at -80°C under optimized and standardized procedures until analysis.

DNA was obtained from children’s peripheral blood (buffy coat) collected in EDTA tubes. DNA was extracted using the Chemagen kit (Perkin Elmer, USA) in batches of 12 samples within each cohort. DNA concentration was determined in a Nanodrop 1000 UV-Vis Spectrophotometer (Thermo Fisher Scientific, USA) and also with Quant-iTTM PicoGreen dsDNA Assay Kit (Life Technologies, USA). DNA extraction was repeated in around 8% of the blood samples as the DNA quantity or quality of the first extraction was low. Less than 1.5% of the samples were finally excluded due to low quality.

RNA was extracted from whole blood samples collected in Tempus tubes (Thermo Fisher Scientific, USA) using MagMAX for Stabilized Blood Tubes RNA Isolation Kit. The quality of RNA was evaluated with a 2100 Bioanalyzer (Agilent Technologies, USA) and the concentration with a NanoDrop 1000 UV-Vis Spectrophotometer. Samples classified as good RNA quality (78.67%) had a similar RNA pattern at visual inspection in the Bioanalyzer, a RNA Integrity Number (RIN) >5 and a concentration >10 ng/ul. Mean values (standard deviation, SD) for the RIN, concentration (ng/ul), Nanodrop 260/280 ratio and Nanodrop 260/230 ratio were: 7.05 (0.72), 109.07 (57.63), 2.15 (0.16) and 0.61 (0.41).

During the clinical examination, two spot urine samples (one before bedtime and one first morning void) were brought by the participants to the research centre in cool packs and stored at 4°C until processing. Urine samples of the night before the visit and the first morning void on the day of the visit were combined to provide Two urine samples, representing last night-time and first morning voids, were collected on the evening and morning before the clinical examination and were subsequently pooled to generate a more representative sample of the last 24 h for metabolomic analysis (n = 1107). Either the night-time void (n = 37) or morning void (n = 48) sample was analysed in cases where a pooled sample was missing [60].

### Telomere length measurement

Blood average relative telomere length was measured by a modified qPCR protocol as described previously [68]. Telomere and single copy-gene reaction mixture and PCR cycles used can be found in Martens et al. [69]. All measurements were performed in triplicate on a 7900HT Fast Real-Time PCR System (Applied Biosystems) in a 384-well format. On each run, a 6-point serial dilution of pooled DNA was run to assess PCR efficiency as well as eight inter-run calibrators to account for the inter-run variability. Relative telomere lengths were calculated using qBase software (Biogazelle, Zwijnaarde, Belgium) and were expressed as the ratio of telomere copy number to single-copy gene number (T/S) relative to the average T/S ratio of the entire sample set. We achieved CV’s within triplicates of the telomere runs, single-copy gene runs, and T/S ratios of 0.84%, 0.43%, and 6.4%, respectively.

### DNA methylation

Blood DNA methylation was assessed with the Infinium HumanMethylatio450 beadchip (Illumina, USA) at the University of Santiago de Compostela – Spanish National Genotyping Center (CeGen-USC) (Spain). 700 ng of DNA were bisulfite-converted using the EZ 96-DNA kit (Zymo Research, USA) following the manufacturer’s standard protocol. All samples of the study were randomized considering sex and cohort. In addition, each plate contained a HapMap control sample and 24 HELIX inter-plate duplicates were included.

After an initial inspection of the quality of the methylation data with the MethylAid package[70], probes with a call rate <95% based on a detection p-value of 1e-16 and samples with a call rate <98% were removed [71]. Samples with discordant sex were eliminated from the study as well as duplicates with inconsistent genotypes and samples with inconsistent genotypes respect to existing genome-wide genotyping array data. Methylation data was normalized using the functional normalization method with prior background correction with Noob [72]. Then, some probes were filtered out: control probes, probes to detect single nucleotide polymorphisms (SNPs), probes to detect methylation in non-CpG sites, probes located in sexual chromosomes, cross hybridizing probes [73], probes containing a SNP at any position of the sequence with a minor allele frequency (MAF) >5% and probes with a SNP at the CpG site or at the single base extension (SBE) at any MAF in the combined population from 1000 Genomes Project. Batch effect (slide) was corrected using the ComBat R package [74]. CpGs were annotated with the IlluminaHumanMethylation450kanno.ilmn12.hg19 R package [75].

### Transcriptome analysis

Gene expression was assessed using the GeneChip® Human Transcriptome Array 2.0 (HTA 2.0) from Affymetrix (USA) at the University of Santiago de Compostela (USC) (Spain). Briefly, RNA samples were concentrated or evaporated in order to reach the required RNA input concentration (200 ng of total RNA). Amplified and biotinylated sense-strand DNA targets were generated from total RNA. Microarrays were hybridized according to the Affymetrix recommendations using the Affymetrix labeling and hybridization kits. All samples were randomized within each batch considering sex and cohort. Two different types of control RNA samples (HeLa and FirstChoice® Human Brain Reference RNA (Thermo Fisher Scientific, USA)) were included in each batch, but they were hybridized only in the first batches.

Raw data were extracted with the Affymetrix AGCC software and normalized with the GCCN (SST-RMA) algorithm at the gene level (http://tools.thermofisher.com/content/sfs/brochures/sst_gccn_whitepaper.pdf). Annotation of transcripts clusters (TCs) to genes was done with the Affymetrix Expression Console software using the HTA-2_0 Transcript Cluster Annotations Release na36 (hg19). A transcript cluster is defined as a group of one or more probes covering a region of the genome reflecting all the exonic transcription evidence known for the region and corresponding to a known or putative gene. Four samples with discordant sex were detected with the MassiR R package [76] and excluded. Control probes, and TCs in sexual chromosomes and without chromosome information were filtered out. Batch effect (slide) was corrected using the ComBat R package [74]. To determine TC call rate, 10 constitutive or best probes based on probe scoring and cross-hybridation potential were selected per TC. Probe Detection Above Background (DABG) p-values were computed based on the rank order against the background probe set intensities. Probe level p-values were combined into a TC level p-value using the Fisher equation. TCs with a DABG p-value <0.05 were defined as detected. Three samples with low call rate (<40%) as well as TCs with a call rate <1% were excluded from the dataset. Gene expression values were log2 transformed.

### Proteome analysis

Plasma protein levels were assessed using the antibody-based multiplexed platform from Luminex. Three kits targeting 43 unique candidate proteins were selected (Thermo Fisher Scientifics, USA): Cytokines 30-plex (Catalog Number (CN): LHC6003M), Apoliprotein 5-plex (CN: LHP0001M) and Adipokine 15-plex (CN: LHC0017M).

All samples were randomized and blocked by cohort prior measurement. For quantification, an 8-point calibration curve per plate was performed with protein standards provided in the Luminex kit and following procedures described by the vendor. Commercial heat inactivated, sterile-filtered plasma from human male AB plasma (Sigma-Aldrich, USA) was used as constant samples to control for intra- and inter-plate variability. Four control samples were added per plate. All samples, including controls, were diluted ½ for the 30-plex kit, ¼ for the 15-plex kit and 1/2500 for the 5-plex kit.

Raw intensities obtained with the xMAP and Luminex system for each plasma sample were converted to pg/ml using the calculated standard curves of each plate and accounting for the dilutions made prior measurement. The percentages of coefficients of variation (CV%) for each protein by plate ranged from 3% to 36%. The limit of detection (LOD) and the lower and upper limit of quantification (LOQ1 and LOQ2, respectively) were estimated by plate, and then averaged. Only proteins with >30% of measurements in the linear range of quantification were kept in the database and the others were removed. Seven proteins were measured twice (in two different multiplex kits). We kept the measure with higher quality. The 36 proteins that passed the quality control criteria mentioned above were log2 transformed [77]. Then, the plate batch effect was corrected by subtracting the plate specific average for each protein minus the overall average of all plates for that protein. After that, values below the LOQ1 and above the LOQ2 were imputed using a truncated normal distribution implemented in the truncdist R package [78]. Twenty samples were excluded due to having ten or more proteins out of the linear range of quantification.

### Metabolomic analysis

The AbsoluteIDQTM p180 kit was chosen for serum analysis as it is a standardised, targeted LC-MS/MS assay, widely used for large-scale epidemiology studies and its inter-laboratory reproducibility has been demonstrated by several independent laboratories [79].Serum samples were quantified using the AbsoluteIDQTM p180 kit following the manufacturer’s protocol (User Manual UM_p180_AB_SCIEX_9, Biocrates Life Sciences AG) using LC-MS/MS; an Agilent HPLC 1100 liquid chromatography coupled to a SCIEX QTRAP 6500 triple quadrupole mass spectrometer. A full description of the HELIX metabolomics methods and data can be found elsewhere [80].

Briefly, the kit allows for the targeted analysis of 188 metabolites in the classes of amino acids, biogenic amines, acylcarnitines, glycerophospholipids, sphingolipids and sum of hexoses, covering a wide range of analytes and metabolic pathways in one targeted assay. The kit consists of a single sample processing procedure, with two separate analytical runs, a combination of liquid chromatography (LC) and flow injection analysis (FIA) coupled to tandem mass spectrometry (MS/MS). Isotopically labelled and chemically homologous internal standards were used for quantification. The AbsoluteIDQ p180 data of serum samples were acquired in 18 batches. Every analytical batch, in a 96-well plate format, included up to 76 randomised cohort samples. Also in every analytical batch, three sets of quality control samples were included, the NIST SRM 1950 plasma reference material (in 4 replicates), a commercial available serum QC material (CQC in 2 replicates, SeraLab, S-123-M-27485) and the QCs provided by the manufacturer in three concentration levels. The NIST SRM 1950 reference was used as the main quality control sample for the LC-MS/MS analysis. Coefficients of variation (CVs) for each metabolite were calculated based on the NIST SRM 1950 and also the limits of detection (LODs) were also used to assess the analytical performance of individual metabolites. Metabolite exclusion was based on a metabolite variable meeting two conditions: (1) CV of over 30% and (2) over 30% of the data are below LOD. Eleven out of the 188 serum metabolites detected were excluded as a result, leaving 177 serum metabolites to be used for further statistical analysis. The mean coefficient of variation across the 177 LC-MS/MS detected serum metabolites was 16%. We also excluded one HELIX sample, which was hemolyzed.

Urinary metabolic profiles were acquired using 1H NMR spectroscopy according to (Lau et al., 2018). In brief one-dimensional 600 MHz 1H NMR spectra of urine samples from each cohort were acquired on the same Bruker Avance III spectrometer operating at 14.1 Tesla within a period of 1 month. The spectrometer was equipped with a Bruker SampleJet system, and a 5-mm broad-band inverse configuration probe maintained at 300K. Prior to analysis, cohort samples were randomised. Deuterated 3-(trimethylsilyl)-[2,2,3,3-d4]-propionic acid sodium salt (TSP) was used as internal reference. Aliquots of the study pooled quality control (QC) sample were used to monitor analytical performance throughout the run and were analysed at an interval of every 23 samples (i.e. 4 QC samples per well plate). The 1H NMR spectra were acquired using a standard one-dimensional solvent suppression pulse sequence. 44 metabolites were identified and quantified as described (Lau et al., 2018). The urinary NMR showed excellent analytical performance, the mean coefficient of variation across the 44 NMR detected urinary metabolites was 11%. Data was normalized using the median fold change normalization method [81], which takes into account the distribution of relative levels of all 44 metabolites compared to the reference sample in determining the most probable dilution factor. An offset of ½ of the minimal value was applied and then concentration levels were expressed as log2.

### Building biological clocks

Child epigenetic age was calculated based on Horvath’s Skin and Blood clock [30] using the methylclock R package [82].

New transcriptome and immunometabolic clocks were trained against chronological age on transcriptome data and concatenated proteomic and metabolomic data respectively, from the HELIX subcohort children through elastic net regression, using the *glmnet* R package [83]. All ‘omic data was first mean centred and univariate scaled. To tune hyperparameters alpha and lambda, we performed a line search for alpha between 0 and 1, in 0.1 increments, and each time found the optimal value of lambda based on minimization of cross-validated mean squared error, using the *cvfits* function and 10-fold cross-validation. The best performing combination of alpha and lambda was reserved for fitting the final model.

Transcriptome data and concatenated proteomic and metabolomic data from the HELIX panel study children, was reserved for testing performance (Pearson’s r and mean absolute error with chronological age) of the derived clocks. Paired, one-tailed t-tests were used to test if biological age measures increased between the HELIX subcohort and subsequent HELIX panel clinical examinations.

### Developmental measurements

During the HELIX subcohort examination, height and weight were measured using regularly calibrated instruments and converted to BMI and height age-and-sex–standardized z-scores (zBMI and zHeight) using the international World Health Organization (WHO) reference curves [84]. Bioelectric impedance analyses were performed with the Bodystat 1500 (Bodystat Ltd.) equipment after 5 min of lying down. The proportion of fat mass was calculated using published age- and race-specific equations validated for use in children [85].

Trained fieldwork technicians measured three cognitive domains in children using a battery of computer-based tests: fluid intelligence (Raven Coloured Progressive Matrices Test [CPM]), attention function (Attention Network Test [ANT]) and working memory (N-Back task). Complete outcome descriptions are provided in [86]. The CPM comprised a total of 36 items and we used the total number of correct responses as the outcome. A higher CPM scoring indicates better fluid intelligence. Fluid intelligence is the ability to solve novel reasoning problems and depends only minimally on prior learning. For ANT, we used the outcome of hit reaction time standard error (HRT-SE), a measure of response speed consistency throughout the test. A high HRT-SE indicates highly variable reaction time during the attention task and is considered a measure of inattentiveness [87]. As the main parameter of N-Back, we used d prime (d′) from the 3-back colours test, a measure derived from signal detection theory calculated by subtracting the z-score of the false alarm rate from the z-score of the hit rate. A higher d′ indicates more accurate test performance, i.e. better working memory [87]. All examiners were previously trained following a standardized assessment protocol by the study expert psychologist. Furthermore, during the pilot phase, a coordinator visited each cohort site and checked for any potential error committed by the previously trained examiners.

Parents completed questionnaires related to child’s behavior, including the Conner rating scale’s (N = 1287) and child behavior checklist (CBCL, N = 1298), within a week before the follow-up visit at 6–11 years of age. The 99-item CBCL/6–18 version for school children was used to obtain standardized parent reports of children’s problem behaviours, translated and validated in each native language of the participating six cohort populations [88]. The parents responded along a 3-point scale with the code of 0 if the item is not true of the child, 1 for sometimes true, and 2 for often true. The internalizing score includes the subscales of emotionally reactive and anxious/depressed symptoms, as well as somatic complaints and symptoms of being withdrawn. The externalizing score includes attention problems and aggressive behaviors.

Lung function was measured by a spirometry test (EasyOne spirometer; NDD [New Diagnostic Design], Zurich, Switzerland), by trained research technicians using a standardised protocol. The child, sitting straight and equipped with a nose clip, was asked to perform at least six manoeuvres (if possible). Details of exclusion of unacceptable maneuvers and validation of acceptable spirometer curves is fully described in [89]. FEV1 percent predicted values were computed using the reference equations estimated by the Global Lung Initiative [90] for computing the (standardised by age, height, sex, and ethnicity).

Parents of children aged 8 years or older completed an additional questionnaire based on the pubertal development scale (PDS) [91]. Boys are asked whether growth has not begun, barely begun, is definitely underway, or has finished on five dimensions: body hair, facial hair, voice change, skin change, and growth spurt. Girls are asked the same questions about body hair, skin change, breast development, and growth spurt. Responses are coded on 4-point scales (1 = no development and 4 = completed development). For girls, a yes-no question about onset of menarche is weighted more heavily (1 = no and 4 = yes). For both genders, ratings are then averaged to create an overall score for physical maturation. Due to the young age of participants, we took the average scores and created a binary variable, to define whether puberty had started (PDS >1) or not (PDS=1).

### Covariates

During pregnancy and in the childhood HELIX subcohort examination information on the following key covariates was collected: cohort study centre (BiB, EDEN, INMA, MoBa, KANC and RHEA), self-reported maternal education (primary school, secondary school and university degree or higher), self-reported ancestry (White European, Asian and Pakistani, or other), birth weight (continuous, kg), gestational age at delivery (continuous in weeks).

Information about the children’s habitual diet was collected via a semi quantitative food-frequency questionnaire (FFQ) covering the child’s habitual diet, which was filled in by the parent attending the examination appointment. The FFQ, covering the past year, was developed by the HELIX research group, translated and applied to all cohorts. For the Mediterranean Diet Quality Index (KIDMED index) [92], items positively associated with the Mediterranean diet pattern (11 items) were assigned a value of +1, while those negatively associated with the Mediterranean diet pattern (4 items) were assigned a value of −1. The scores for all 15 items were summed, resulting in a total KIDMED score ranging from −4 to 11, with higher scores reflecting greater adherence to a Mediterranean diet.

The smoking status of the mother at any point during pregnancy was categorised into “non-active smoker, or “active smoker”. Global exposure of the child to environmental tobacco smoke was defined based on the questionnaires completed by the parents into: “no exposure”, no exposure at home neither in other places; “exposure”: exposure in at least one place, at home or outside.

Moderate-to-vigorous physical activity variable was created based on physical activity questionnaire developed by the HELIX research group. It was defined as the amount of time children spent doing physical activities with intensity above 3 metabolic equivalent tasks (METs) and is expressed in units of min/day.

Family Affluence Score (FAS) [93]was included based on questions from the subcohort questionnaire. A composite FAS score was calculated based on the responses to the next four items: (1) Does your family own a car, van or truck? (2) Do you have your own bedroom for yourself? (3) During the past 12 months, how many times did you travel away on holiday with your family? (4) How many computers does your family own? A three-point ordinal scale was used, where FAS low (score 0,1,2) indicates low affluence, FAS medium (score 3,4,5) indicates middle affluence, and FAS high (score 6,7,8,9) indicates high affluence FAS.

Family social capital-related questions were included in the HELIX questionnaire to capture different aspects of social capital, relating both to the cognitive (feelings about relationships) and structural (number of friends, number of organizations) dimensions and to bonding capital (close friends and family), bridging capital (neighbourhood connections, looser ties) and linking capital (ties across power levels; for example, political membership). Family social capital was categorized into low, medium and high based on terciles.

### Statistical analysis

All statistical analyses described here were performed among the HELIX subcohort children only. Since there were few missing covariate data (table S6), complete-case analysis was performed. Correlations between biological age measures and chronological age were calculated using Pearson’s correlations. Partial correlations, adjusted for chronological age and cohort study centre, were applied to assess correlations between biological age measures.

In analysis with health risk factors and developmental outcomes, relative telomere length was multiplied by -1 to provide directions of effect consistent with the biological age clocks and univariate scaled to express effects in terms of SD change in telomere length. The markers derived from omic-based biological clocks were expressed as Δ age (clock-predicted age – chronological age). Associations between the biological age markers and developmental measures were estimated using linear regression, or logistic regression for onset of puberty, with the developmental measure as the dependent variable. CBCL scores were log transformed to achieve an approximately normal distribution. Continuous outcomes, apart from the BMI and height z-scores, were mean centered and univariate scaled for the purposes of graphical representation. Associations between health risk factors and biological age markers were estimated using linear regression with the biological age marker as the dependent variable. All regression analyses were adjusted for chronological age, sex, ethnicity, and study centre.

We preformed three sensitivity analyses: Firstly, we repeated analysis with health outcomes stratified by child sex, since the relationship between biological age and development may differ between boys and girls. Secondly, we further adjusted regression models for estimated cell counts (CD4T, CD8T, monocytes, B cells, NK cells, neutrophils and eosinophils), since it has been proposed for epigenetic clocks that cell proportion adjustments allow estimation of effects on the intrinsic cellular ageing rate, rather than the extrinsic rate outputted by blood based biological clocks, which may be partly determined by age-related changes in cell composition [94]. Blood cell type proportion was estimated from DNA methylation data using the Reinius et al. [79] reference panel as implemented in meffil package [80]. Finally, we assessed the effects of further adjustment for health risk factors identified as associated with any of the biological age markers (family affluence and social capital, birthweight, maternal active smoking, and child passive smoking). In our main analysis, we have not adjusted for these factors as our assumption is that the effects of health risk factors on child development is mediated through biological age. However, an alternative assumption is that health risk factors exert independent effects on both biological age and developmental outcomes, which would require adjustment for these factors to estimate direct effects of biological age on developmental outcomes.

We report associations significant at the both the nominal significance threshold (p <0.05) and after correction for 5% false discovery rate using the Benjamini and Hochberg [95] method, calculated across all computed associations.

We performed overrepresentation analyses (ORA) among KEGG and REACTOME pathways and gene ontology (GO) sets of all transcripts contributing to the transcriptome clock using the ConsensuspathDB online tool (http://consensuspathdb.org/). A pathway or GO set was considered significantly enriched if FDR corrected p-values were smaller than 0⋅05 and included at least 3 genes. Additionally, to assess concordance with gene expression changes with age in adults, we tested enrichment of all transcripts contributing to the transcriptome clock among age-associated transcripts reported by Peters et al. [8], using a hypergeometric test using the R “phyper” function.

All analyses were performed in R version 4.1.2.

## Data Availability

Due to data protection regulations in each participating country and participant data use agreements, human subject data used in this project cannot be freely shared. The raw data supporting the current study are available on request subject to ethical and legislative review. The "HELIX Data External Data Request Procedures" are available with the data inventory in this website: http://www.projecthelix.eu/data-inventory. The document describes who can apply to the data and how, the timings for approval and the conditions to data access and publication. Researchers who have an interest in using data from this project for reproducibility or in using data held in general in the HELIX data warehouse for research purposes can apply for access to data. Interested researchers should fill in the application protocol found in ANNEX I at https://www.projecthelix.eu/files/helix_external_data_request_procedures_final.pdf and send this protocol to. The applications are received by the HELIX Coordinator, and are processed and approved by the HELIX Project Executive Committee. All code used for data analysis has been provided as supplementary material.

## Data availability

Due to data protection regulations in each participating country and participant data use agreements, human subject data used in this project cannot be freely shared. The raw data supporting the current study are available on request subject to ethical and legislative review. The “HELIX Data External Data Request Procedures” are available with the data inventory in this website: http://www.projecthelix.eu/data-inventory. The document describes who can apply to the data and how, the timings for approval and the conditions to data access and publication. Researchers who have an interest in using data from this project for reproducibility or in using data held in general in the HELIX data warehouse for research purposes can apply for access to data. Interested researchers should fill in the application protocol found in ANNEX I at https://www.projecthelix.eu/files/helix_external_data_request_procedures_final.pdf and send this protocol to. The applications are received by the HELIX Coordinator, and are processed and approved by the HELIX Project Executive Committee. All code used for data analysis has been provided as supplementary material. Deidentified dataset for generation of figures 1 and 2 has been provided as a supplementary dataset.

## Acknowledgments

The authors are grateful to all the participating families in the six countries who took part in this study and to all the field workers for their dedication and efficiency. ISGlobal acknowledges support from the Spanish Ministry of Science and Innovation through the “Centro de Excelencia Severo Ochoa 2019–2023” Program (CEX 2018-000806-S), and support from the Generalitat de Catalunya through the CERCA Programme. The CRG/UPF Proteomics Unit is part of the Spanish Infrastructure for Omics Technologies (ICTS OmicsTech) and it is supported by “Secretaria d’Universitats i Recerca del Departament d’Economia i Coneixement de la Generalitat de Catalunya” (2017SGR595). We also acknowledge support of the Spanish Ministry of Science and Innovation to the EMBL partnership, the Centro de Excelencia Severo Ochoa and the CERCA Programme / Generalitat de Catalunya. Some figures were created with BioRender.com

## Supplementary Figures

**Figure s1:**
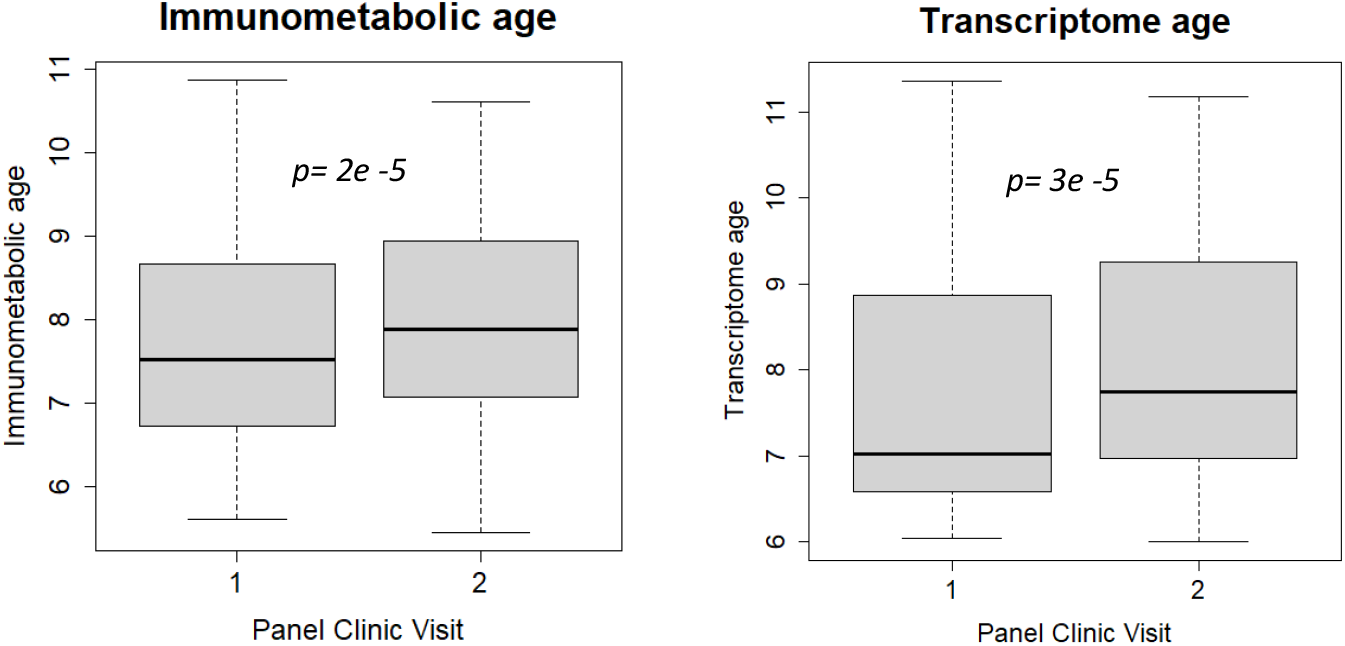
Comparison between immunometabolic and transcriptome age between first and second study visits. Box plots (showing minimum, maximum, median, first quartile and third quartile) of biological age measures at each panel study visit (approximately 6 months apart). Panel clinic 1 was part of the main Helix subcohort examination. P values calculated from paired t-tests.

**Figure s2:**
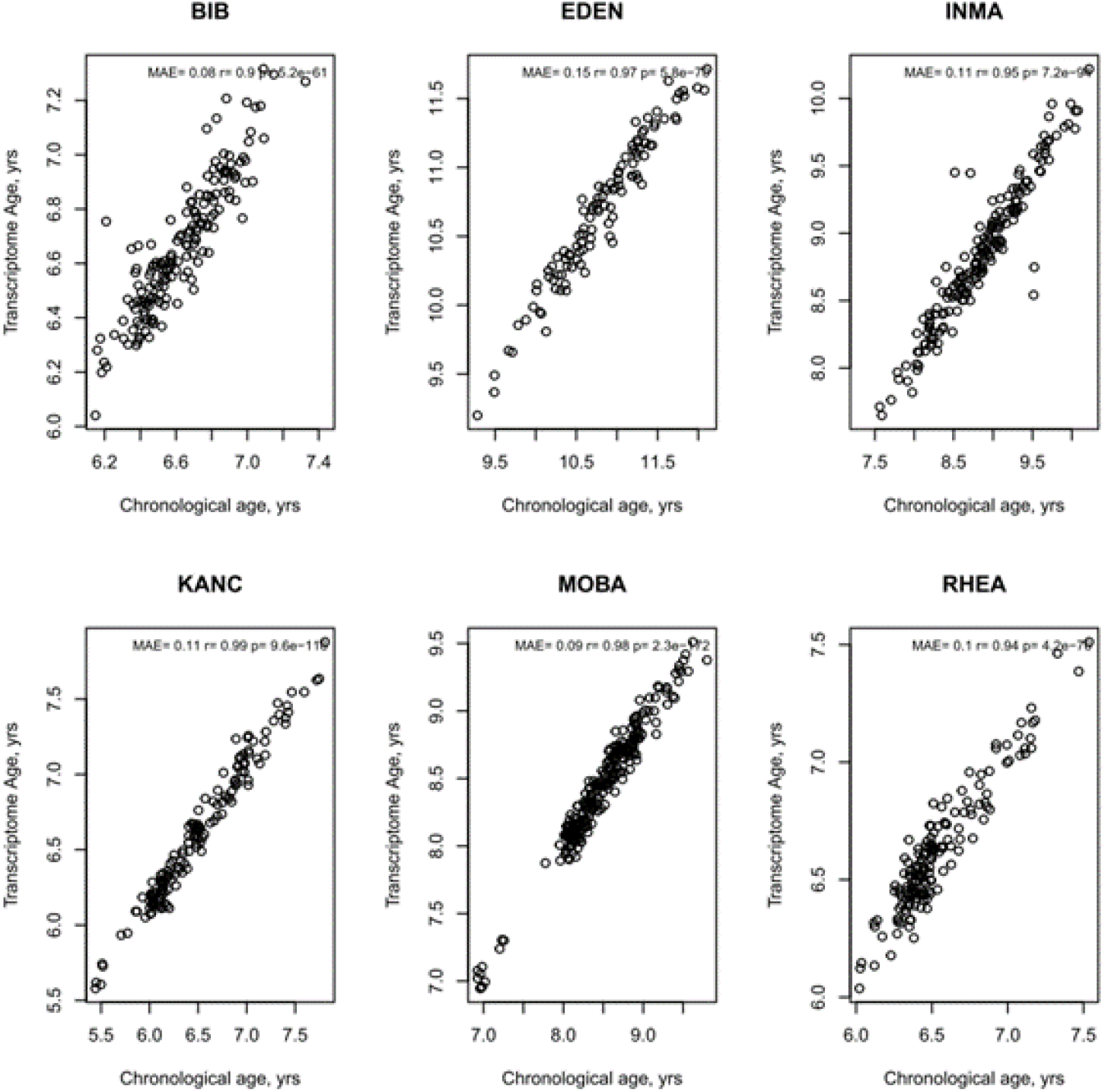
Age Prediction by study centre of transcriptome age. MAE = mean absolute error. *r* and *p* values from Pearson’s correlation.

**Figure s3:**
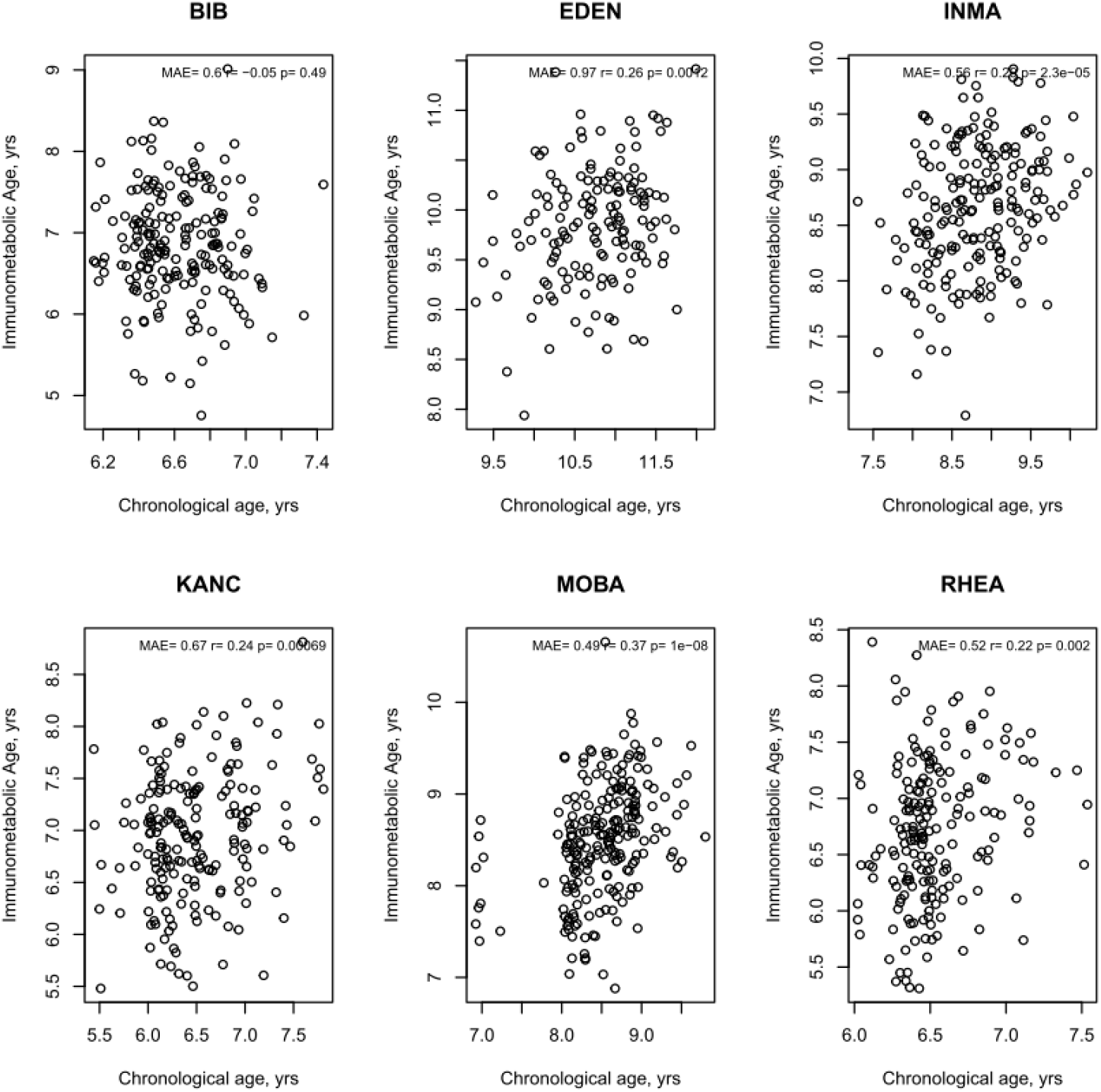
Age Prediction by study centre of immunometabolic age. MAE = mean absolute error. *r* and *p* values from Pearson’s correlation.

**Figure s4:**
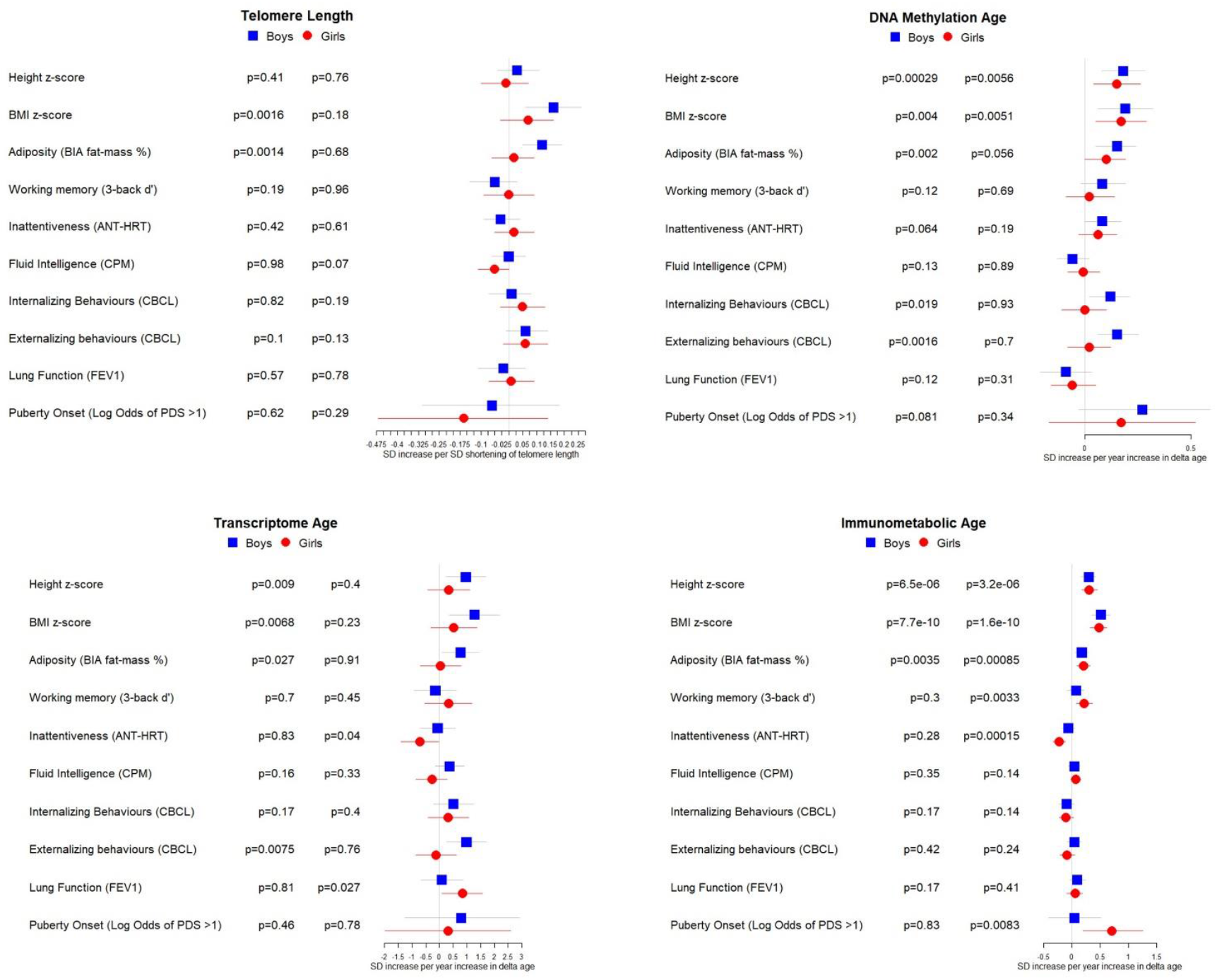
Associations between biological age measures and developmental measures, stratified by sex. Estimates calculated using linear regression, adjusted for chronological age, sex, ethnicity, and study centre. Telomere length is expressed as % decrease in length (multiplied by -1) to provide estimates indicative of accelerated biological age, as for the other biological age indicators.

**Figure s5:**
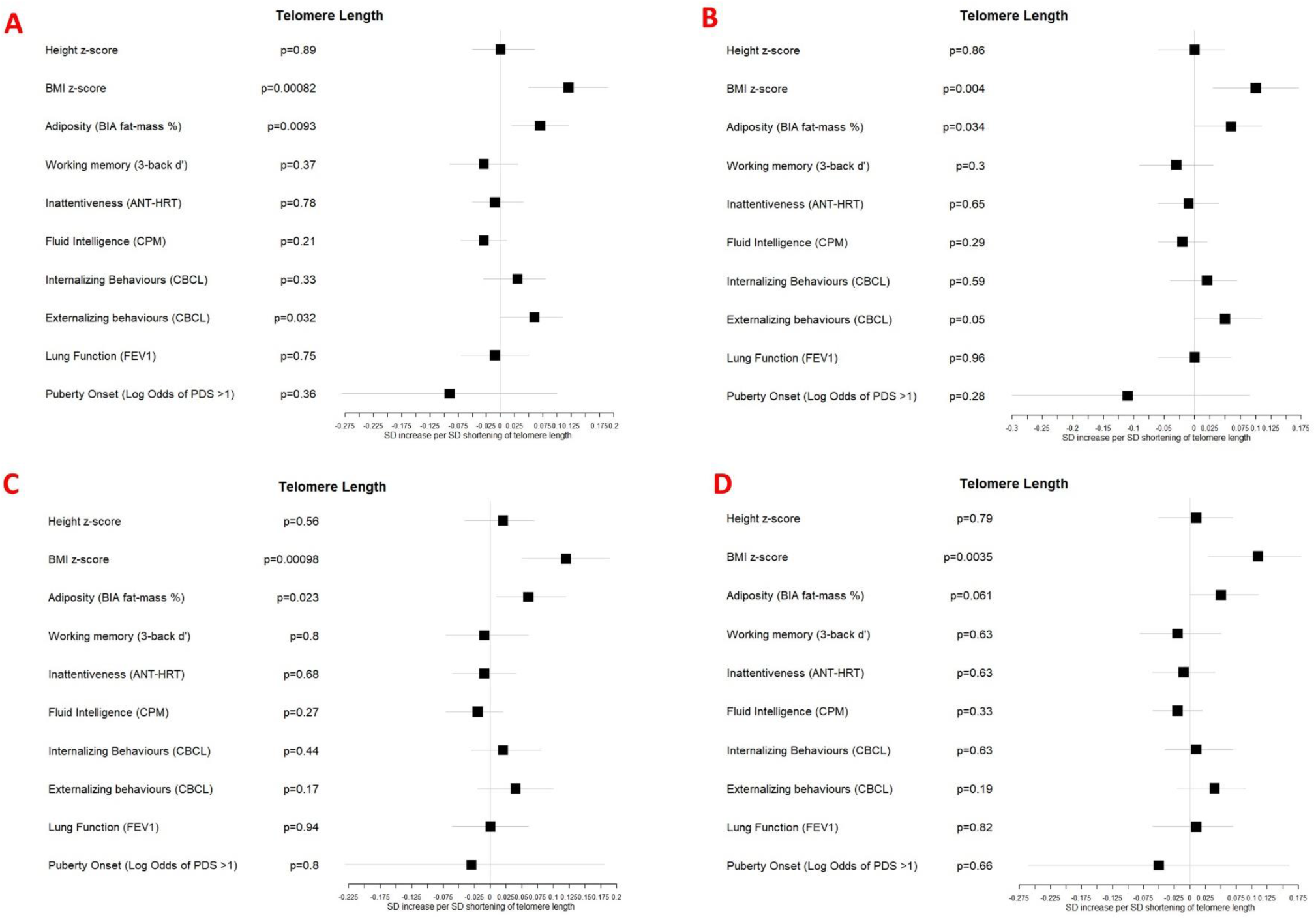
Associations between telomere length and developmental measures adjusted for A: chronological age, sex, ethnicity, and study centre; B: as for A plus estimated cell counts; C: as for A plus family affluence and social capital, birthweight, maternal active smoking, and child passive smoking; D as for C plus estimated cell counts. Telomere length is expressed as % decrease in length (multiplied by -1) to provide estimates indicative of accelerated biological age, as for the other biological age indicators.

**Figure s6:**
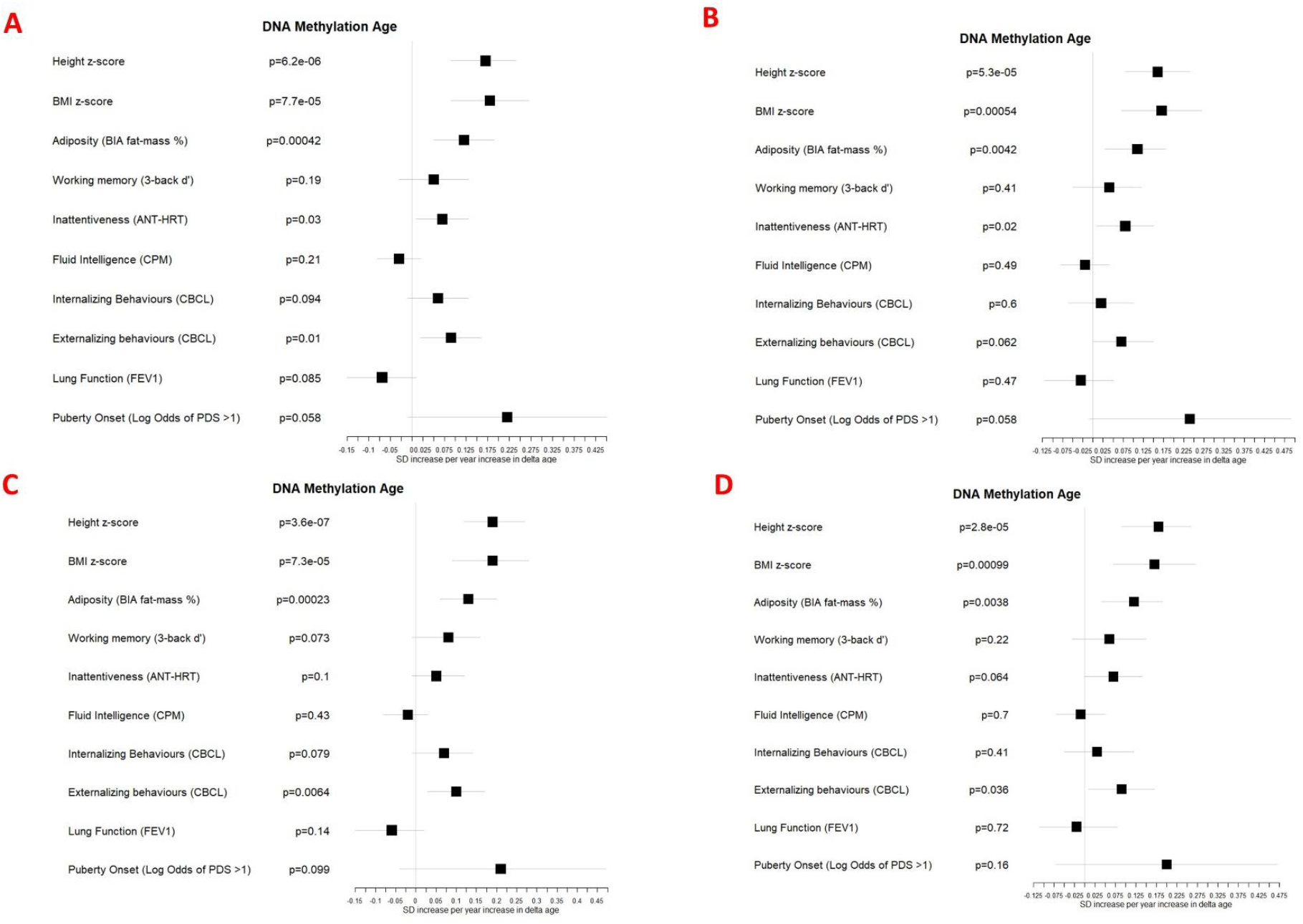
Associations between DNA methylation Δ age and developmental measures adjusted for A: chronological age, sex, ethnicity, and study centre; B: as for A plus estimated cell counts; C: as for A plus family affluence and social capital, birthweight, maternal active smoking, and child passive smoking; D as for C plus estimated cell counts.

**Figure s7:**
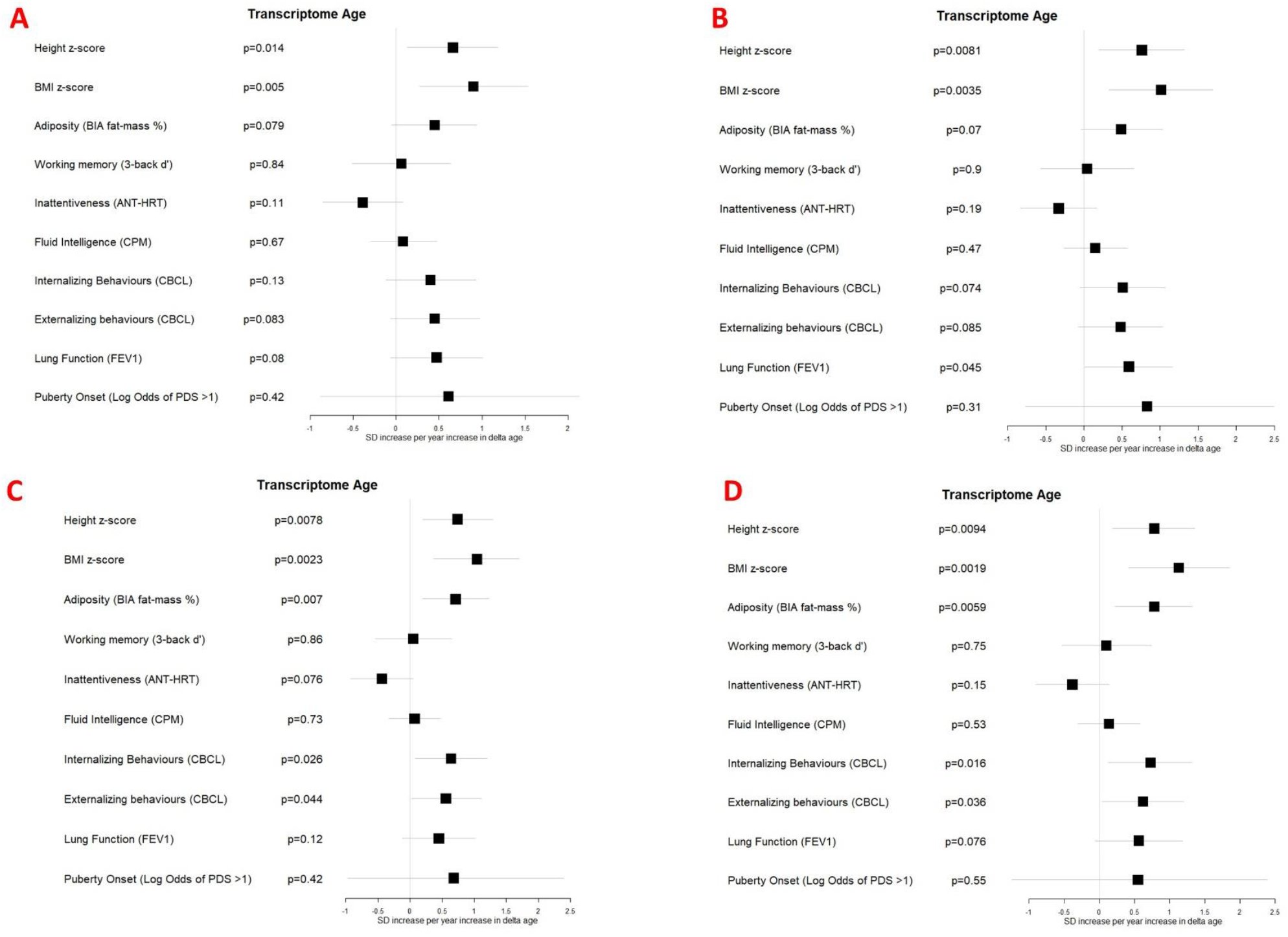
Associations between transcriptome Δ age and developmental measures adjusted for A: chronological age, sex, ethnicity, and study centre; B: as for A plus estimated cell counts; C: as for A plus family affluence and social capital, birthweight, maternal active smoking, and child passive smoking; D as for C plus estimated cell counts.

**Figure s8:**
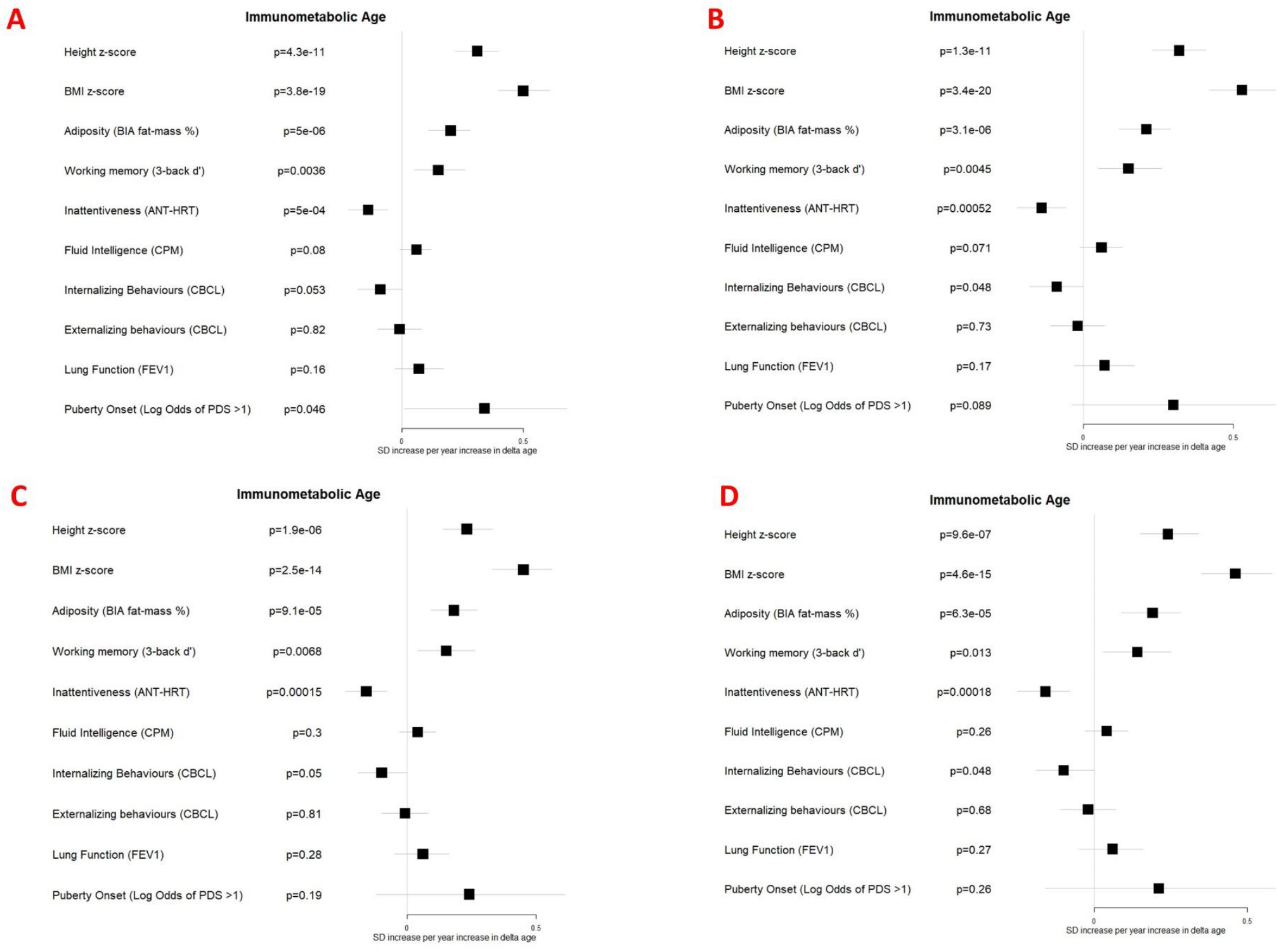
Associations between immunometabolic Δ age and developmental measures adjusted for A: chronological age, sex, ethnicity, and study centre; B: as for A plus estimated cell counts; C: as for A plus family affluence and social capital, birthweight, maternal active smoking, and child passive smoking; D as for C plus estimated cell count

**Figure s9:**
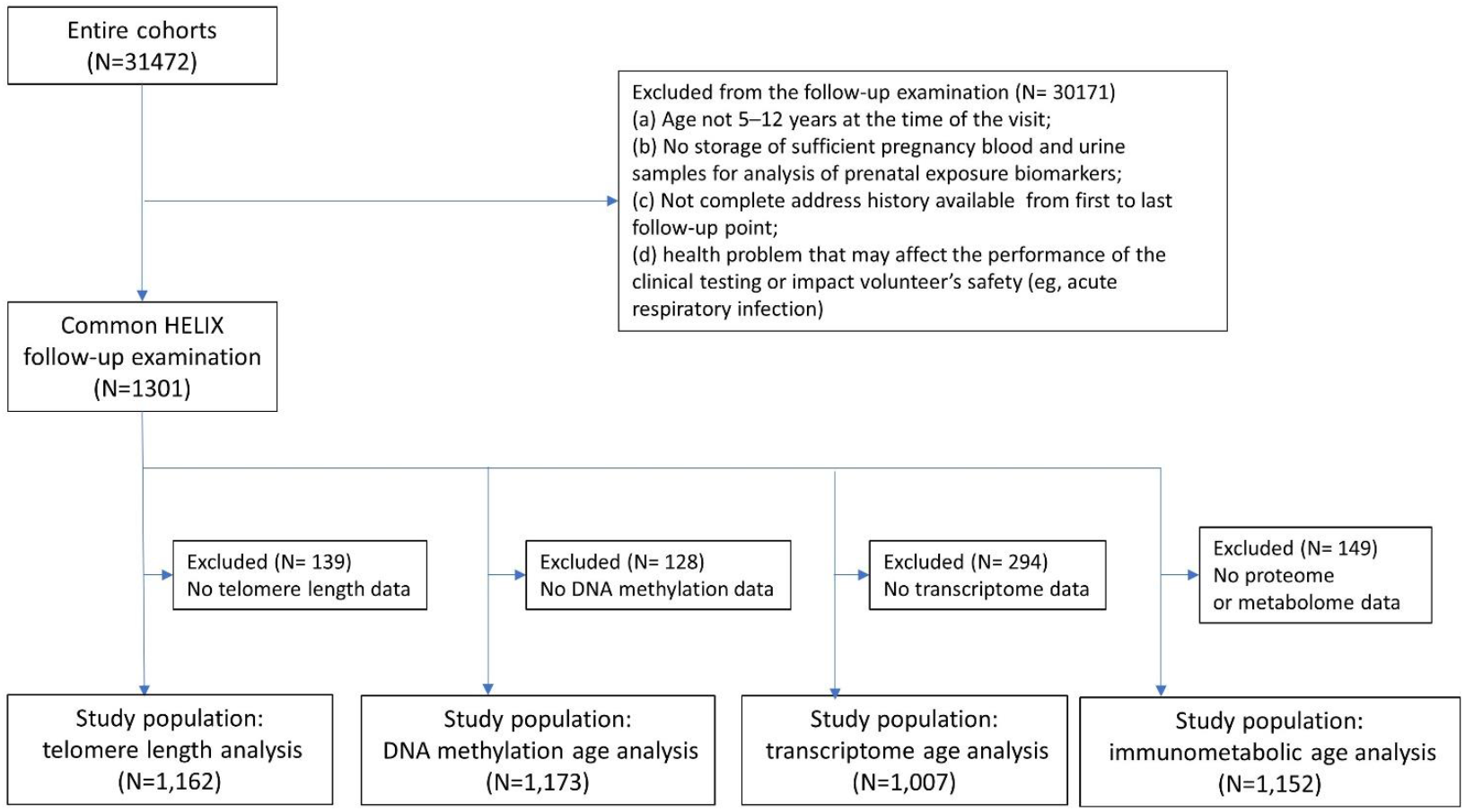
Participant flowchart

## References

1. Sierra, F., The Emergence of Geroscience as an Interdisciplinary Approach to the Enhancement of Health Span and Life Span. Cold Spring Harb Perspect Med, 2016. 6(4): p. a025163.

2. López-Otín, C., et al., The hallmarks of aging. Cell, 2013. 153(6): p. 1194–217.

3. Jylhävä, J., N.L. Pedersen, and S. Hägg, Biological Age Predictors. EBioMedicine, 2017. 21: p. 29–36.

4. Rutledge, J., H. Oh, and T. Wyss-Coray, Measuring biological age using omics data. Nat Rev Genet, 2022.

5. Hertel, J., et al., Measuring Biological Age via Metabonomics: The Metabolic Age Score. J Proteome Res, 2016. 15(2): p. 400–10.

6. Perna, L., et al., Epigenetic age acceleration predicts cancer, cardiovascular, and all-cause mortality in a German case cohort. Clin Epigenetics, 2016. 8: p. 64.

7. Marioni, R.E., et al., The epigenetic clock is correlated with physical and cognitive fitness in the Lothian Birth Cohort 1936. Int J Epidemiol, 2015. 44(4): p. 1388–96.

8. Peters, M.J., et al., The transcriptional landscape of age in human peripheral blood. Nat Commun, 2015. 6: p. 8570.

9. Lehallier, B., et al., Undulating changes in human plasma proteome profiles across the lifespan. Nature Medicine, 2019. 25(12): p. 1843–1850.

10. van den Akker, E.B., et al., Metabolic Age Based on the BBMRI-NL (1)H-NMR Metabolomics Repository as Biomarker of Age-related Disease. Circ Genom Precis Med, 2020. 13(5): p. 541–547.

11. Macdonald-Dunlop, E., et al., A catalogue of omics biological ageing clocks reveals substantial commonality and associations with disease risk. Aging (Albany NY), 2022. 14(2): p. 623–659.

12. Horvath, S., DNA methylation age of human tissues and cell types. Genome Biol, 2013. 14(10): p. R115.

13. Lu, A.T., et al., DNA methylation GrimAge strongly predicts lifespan and healthspan. Aging (Albany NY), 2019. 11(2): p. 303–327.

14. Belsky, D.W., et al., Quantification of the pace of biological aging in humans through a blood test, the DunedinPoAm DNA methylation algorithm. Elife, 2020. 9.

15. Robinson, O., et al., Determinants of accelerated metabolomic and epigenetic aging in a UK cohort. Aging Cell, 2020. 19(6): p. e13149.

16. Sayed, N., et al., An inflammatory aging clock (iAge) based on deep learning tracks multimorbidity, immunosenescence, frailty and cardiovascular aging. Nature Aging, 2021. 1(7): p. 598–615.

17. Belsky, D.W., et al., Eleven Telomere, Epigenetic Clock, and Biomarker-Composite Quantifications of Biological Aging: Do They Measure the Same Thing? Am J Epidemiol, 2018. 187(6): p. 1220–1230.

18. Jansen, R., et al., An integrative study of five biological clocks in somatic and mental health. Elife, 2021. 10.

19. Kuh, D., A life course approach to healthy aging, frailty, and capability. J Gerontol A Biol Sci Med Sci, 2007. 62(7): p. 717–21.

20. Emery Thompson, M., Evolutionary Approaches in Aging Research. Cold Spring Harb Perspect Med, 2022.

21. Horvath, S. and K. Raj, DNA methylation-based biomarkers and the epigenetic clock theory of ageing. Nat Rev Genet, 2018. 19(6): p. 371–384.

22. McEwen, L.M., et al., The PedBE clock accurately estimates DNA methylation age in pediatric buccal cells. Proc Natl Acad Sci U S A, 2020. 117(38): p. 23329–23335.

23. Ferrucci, L., et al., Measuring biological aging in humans: A quest. Aging Cell, 2020. 19(2): p. e13080.

24. Binder, A.M., et al., Faster ticking rate of the epigenetic clock is associated with faster pubertal development in girls. Epigenetics, 2018. 13(1): p. 85–94.

25. Simpkin, A.J., et al., The epigenetic clock and physical development during childhood and adolescence: longitudinal analysis from a UK birth cohort. Int J Epidemiol, 2017. 46(2): p. 549–558.

26. Suarez, A., et al., The epigenetic clock and pubertal, neuroendocrine, psychiatric, and cognitive outcomes in adolescents. Clin Epigenetics, 2018. 10(1): p. 96.

27. Martens, D.S., et al., Newborn telomere length predicts later life telomere length: Tracking telomere length from birth to child- and adulthood. eBioMedicine, 2021. 63.

28. Coimbra, B.M., et al., Stress-related telomere length in children: A systematic review. J Psychiatr Res, 2017. 92: p. 47–54.

29. Giallourou, N., et al., Metabolic maturation in the first 2 years of life in resource-constrained settings and its association with postnatal growths. Sci Adv, 2020. 6(15): p. eaay5969.

30. Horvath, S., et al., Epigenetic clock for skin and blood cells applied to Hutchinson Gilford Progeria Syndrome and <i>ex vivo</i> studies. Aging, 2018. 10(7): p. 1758–1775.

31. de Prado-Bert, P., et al., The early-life exposome and epigenetic age acceleration in children. Environ Int, 2021. 155: p. 106683.

32. Hotamisligil, G.S., Inflammation, metaflammation and immunometabolic disorders. Nature, 2017. 542(7640): p. 177–185.

33. Nevalainen, T., et al., Obesity accelerates epigenetic aging in middle-aged but not in elderly individuals. Clin Epigenetics, 2017. 9: p. 20.

34. Fiorito, G., et al., Socioeconomic position, lifestyle habits and biomarkers of epigenetic aging: a multi-cohort analysis. Aging (Albany NY), 2019. 11(7): p. 2045–2070.

35. López-Otín, C., et al., Metabolic Control of Longevity. Cell, 2016. 166(4): p. 802–821.

36. Suzuki, K., et al., Relationship between obesity and serum markers of oxidative stress and inflammation in Japanese. Asian Pac J Cancer Prev, 2003. 4(3): p. 259–66.

37. Minamino, T., et al., A crucial role for adipose tissue p53 in the regulation of insulin resistance. Nat Med, 2009. 15(9): p. 1082–7.

38. Davey Smith, G., et al., Height and risk of death among men and women: aetiological implications of associations with cardiorespiratory disease and cancer mortality. J Epidemiol Community Health, 2000. 54(2): p. 97–103.

39. Tanisawa, K., et al., Inverse Association Between Height-Increasing Alleles and Extreme Longevity in Japanese Women. The Journals of Gerontology: Series A, 2017. 73(5): p. 588–595.

40. Li, Q., et al., Dose–response association between adult height and all-cause mortality: a systematic review and meta-analysis of cohort studies. European Journal of Public Health, 2020. 31(3): p. 652–658.

41. Huang, S.-Y., et al., Investigating causal relationships between exposome and human longevity: a Mendelian randomization analysis. BMC Medicine, 2021. 19(1): p. 150.

42. He, Q., et al., Shorter Men Live Longer: Association of Height with Longevity and FOXO3 Genotype in American Men of Japanese Ancestry. PLOS ONE, 2014. 9(5): p. e94385.

43. Costa Dde, S., et al., Telomere length is highly inherited and associated with hyperactivity-impulsivity in children with attention deficit/hyperactivity disorder. Front Mol Neurosci, 2015. 8: p. 28.

44. Daoust, A.R., et al., Associations Between Children’s Telomere Length, Parental Intrusiveness, and the Development of Early Externalizing Behaviors. Child Psychiatry Hum Dev, 2021.

45. Wojcicki, J.M., et al., Telomere length is associated with oppositional defiant behavior and maternal clinical depression in Latino preschool children. Transl Psychiatry, 2015. 5(6): p. e581.

46. Kroenke, C.H., et al., Autonomic and adrenocortical reactivity and buccal cell telomere length in kindergarten children. Psychosom Med, 2011. 73(7): p. 533–40.

47. Bourdieu, P., The forms of capital, in Hand-book of theory and research for the sociology of education, R. J, Editor. 1986, Greenwood: New York. p. 241–58.

48. Vineis, P. and M. Kelly-Irving, Biography and biological capital. Eur J Epidemiol, 2019. 34(10): p. 979–982.

49. Gardner, M., et al., Gender and telomere length: systematic review and meta-analysis. Exp Gerontol, 2014. 51: p. 15–27.

50. Buxton, J.L., et al., Childhood obesity is associated with shorter leukocyte telomere length. J Clin Endocrinol Metab, 2011. 96(5): p. 1500–5.

51. Ly, K., et al., Telomere length in early childhood is associated with sex and ethnicity. Sci Rep, 2019. 9(1): p. 10359.

52. Okuda, K., et al., Telomere length in the newborn. Pediatr Res, 2002. 52(3): p. 377–81.

53. León, I., J.M. Cimadevilla, and L. Tascón, Developmental gender differences in children in a virtual spatial memory task. Neuropsychology, 2014. 28(4): p. 485–95.

54. Hägg, S. and J. Jylhävä, Sex differences in biological aging with a focus on human studies. Elife, 2021. 10.

55. Koss, K.J., et al., Early Puberty and Telomere Length in Preadolescent Girls and Mothers. J Pediatr, 2020. 222: p. 193-199.e5.

56. Cancer”, C.G.o.H.F.i.B., Menarche, menopause, and breast cancer risk: individual participant meta-analysis, including 118 964 women with breast cancer from 117 epidemiological studies. Lancet Oncol, 2012. 13(11): p. 1141–51.

57. Charalampopoulos, D., et al., Age at menarche and risks of all-cause and cardiovascular death: a systematic review and meta-analysis. Am J Epidemiol, 2014. 180(1): p. 29–40.

58. Zhang, Q., et al., Improved precision of epigenetic clock estimates across tissues and its implication for biological ageing. Genome Med, 2019. 11(1): p. 54.

59. Pontzer, H., et al., Daily energy expenditure through the human life course. Science, 2021. 373(6556): p. 808–812.

60. Maitre, L., et al., Human Early Life Exposome (HELIX) study: a European population-based exposome cohort. BMJ Open, 2018. 8(9): p. e021311.

61. Vrijheid, M., et al., The human early-life exposome (HELIX): project rationale and design. Environ Health Perspect, 2014. 122(6): p. 535–44.

62. Wright, J., et al., Cohort Profile: the Born in Bradford multi-ethnic family cohort study. Int J Epidemiol, 2013. 42(4): p. 978–91.

63. Heude, B., et al., Cohort Profile: The EDEN mother-child cohort on the prenatal and early postnatal determinants of child health and development. Int J Epidemiol, 2016. 45(2): p. 353–63.

64. Guxens, M., et al., Cohort Profile: the INMA--INfancia y Medio Ambiente--(Environment and Childhood) Project. Int J Epidemiol, 2012. 41(4): p. 930–40.

65. Grazuleviciene, R., et al., Maternal smoking, GSTM1 and GSTT1 polymorphism and susceptibility to adverse pregnancy outcomes. Int J Environ Res Public Health, 2009. 6(3): p. 1282–97.

66. Magnus, P., et al., Cohort Profile Update: The Norwegian Mother and Child Cohort Study (MoBa). Int J Epidemiol, 2016. 45(2): p. 382–8.

67. Chatzi, L., et al., Cohort Profile: The Mother-Child Cohort in Crete, Greece (Rhea Study). Int J Epidemiol, 2017. 46(5): p. 1392–1393k.

68. Cawthon, R.M., Telomere length measurement by a novel monochrome multiplex quantitative PCR method. Nucleic Acids Res, 2009. 37(3): p. e21.

69. Martens, D.S., et al., Maternal pre-pregnancy body mass index and newborn telomere length. BMC Med, 2016. 14(1): p. 148.

70. van Iterson, M., et al., MethylAid: visual and interactive quality control of large Illumina 450k datasets. Bioinformatics, 2014. 30(23): p. 3435–7.

71. Lehne, B., et al., A coherent approach for analysis of the Illumina HumanMethylation450 BeadChip improves data quality and performance in epigenome-wide association studies. Genome Biol, 2015. 16(1): p. 37.

72. Fortin, J.P., et al., Functional normalization of 450k methylation array data improves replication in large cancer studies. Genome Biol, 2014. 15(12): p. 503.

73. Chen, Y.A., et al., Discovery of cross-reactive probes and polymorphic CpGs in the Illumina Infinium HumanMethylation450 microarray. Epigenetics, 2013. 8(2): p. 203–9.

74. Johnson, W.E., C. Li, and A. Rabinovic, Adjusting batch effects in microarray expression data using empirical Bayes methods. Biostatistics, 2006. 8(1): p. 118–127.

75. Hansen KD and A. M, IlluminaHumanMethylation450kmanifest: Annotation for Illumina’s 450k methylation arrays.. 2012.

76. Buckberry, S., et al., massiR : a method for predicting the sex of samples in gene expression microarray datasets. Bioinformatics, 2014. 30(14): p. 2084–2085.

77. Vives-Usano, M., et al., In utero and childhood exposure to tobacco smoke and multi-layer molecular signatures in children. BMC Med, 2020. 18(1): p. 243.

78. Nadarajah, S. and S. Kotz, The Exponentiated Type Distributions. Acta Applicandae Mathematica, 2006. 92(2): p. 97–111.

79. Siskos, A.P., et al., Interlaboratory Reproducibility of a Targeted Metabolomics Platform for Analysis of Human Serum and Plasma. Anal Chem, 2017. 89(1): p. 656–665.

80. Lau, C.E., et al., Determinants of the urinary and serum metabolome in children from six European populations. BMC Med, 2018. 16(1): p. 202.

81. Dieterle, F., et al., Probabilistic quotient normalization as robust method to account for dilution of complex biological mixtures. Application in 1H NMR metabonomics. Anal Chem, 2006. 78(13): p. 4281–90.

82. Pelegí-Sisó, D., et al., methylclock: a Bioconductor package to estimate DNA methylation age. Bioinformatics, 2021. 37(12): p. 1759–1760.

83. Zou, H. and T. Hastie, Regularization and variable selection via the elastic net. Journal of the Royal Statistical Society: Series B (Statistical Methodology), 2005. 67(2): p. 301–320.

84. de Onis, M., et al., Development of a WHO growth reference for school-aged children and adolescents. Bull World Health Organ, 2007. 85(9): p. 660–7.

85. Clasey, J.L., et al., A new BIA equation estimating the body composition of young children. Obesity (Silver Spring), 2011. 19(9): p. 1813–7.

86. Julvez, J., et al., Early life multiple exposures and child cognitive function: A multi-centric birth cohort study in six European countries. Environ Pollut, 2021. 284: p. 117404.

87. Forns, J., et al., The n-back test and the attentional network task as measures of child neuropsychological development in epidemiological studies. Neuropsychology, 2014. 28(4): p. 519–529.

88. Achenbach, T.M., Manual for ASEBA school-age forms & profiles. University of Vermont, Research Center for Children, Youth & Families, 2001.

89. Agier, L., et al., Early-life exposome and lung function in children in Europe: an analysis of data from the longitudinal, population-based HELIX cohort. Lancet Planet Health, 2019. 3(2): p. e81–e92.

90. Quanjer, P.H., et al., Multi-ethnic reference values for spirometry for the 3–95-yr age range: the global lung function 2012 equations. 2012, Eur Respiratory Soc.

91. Petersen, A.C., et al., A self-report measure of pubertal status: Reliability, validity, and initial norms. J Youth Adolesc, 1988. 17(2): p. 117–33.

92. Serra-Majem, L., et al., Food, youth and the Mediterranean diet in Spain. Development of KIDMED, Mediterranean Diet Quality Index in children and adolescents. Public Health Nutr, 2004. 7(7): p. 931–5.

93. Boyce, W., et al., The family affluence scale as a measure of national wealth: validation of an adolescent self-report measure. Social indicators research, 2006. 78(3): p. 473–487.

94. Chen, B.H., et al., DNA methylation-based measures of biological age: meta-analysis predicting time to death. Aging, 2016. 8(9): p. 1844–1865.

95. Benjamini, Y. and Y. Hochberg, Controlling the false discovery rate: a practical and powerful approach to multiple testing. Journal of the Royal statistical society: series B (Methodological), 1995. 57(1): p. 289–300.

